# Electrocardiographic Manifestations of Immune Checkpoint Inhibitor Myocarditis

**DOI:** 10.1101/2021.02.28.21252516

**Authors:** John R Power, Joachim Alexandre, Arrush Choudhary, Benay Ozbay, Salim Hayek, Aarti Asnani, Yuichi Tamura, Mandar Aras, Jennifer Cautela, Franck Thuny, Lauren Gilstrap, Dimitri Arangalage, Steven Ewer, Shi Huang, Anita Deswal, Nicolas L. Palaskas, Daniel Finke, Lorenz Lehman, Stephane Ederhy, Javid Moslehi, Joe-Elie Salem

## Abstract

**Importance:** Immune-checkpoint inhibitor (ICI)-myocarditis often presents with arrhythmias, but electrocardiographic (ECG) findings have not been well described. ICI-myocarditis and acute cellular rejection (ACR) following cardiac transplantation share similarities on histopathology; however, whether they differ in arrhythmogenicity is unclear.

**Objectives:** To describe ECG findings in ICI-myocarditis, compare them to ACR, and evaluate their prognostic significance.

**Design:** Cases of ICI-myocarditis were retrospectively identified through a multicenter network. Grade 2R or 3R ACR was retrospectively identified within one center. Two blinded cardiologists interpreted ECGs.

**Setting:** 49 medical centers spanning 11 countries.

**Participants:** 147 adults with ICI-myocarditis, 50 adults with ACR.

**Exposure:** Myocarditis after ICI exposure per European Society of Cardiology criteria for clinically suspected myocarditis, grade 2R or 3R ACR per the International Society for Heart and Lung Transplantation working formulation for biopsy diagnosis of rejection.

**Outcomes:** All-cause mortality, myocarditis-related mortality; and composite endpoint (defined as myocarditis-related mortality and life-threatening ventricular arrhythmia).

**Results:** Of 147 patients, the median age was 67 years (58-77) with 92 (62.6%) men. At 30 days, ICI-myocarditis had an all-cause mortality of 39/146(26.7%), myocarditis-related mortality of 24/146(16.4%), and composite endpoint of 37/146(25.3%). All-cause mortality was more common in patients who developed complete heart block (12/25[48%] vs 27/121[22.3%], hazard ratio (HR)=2.62, 95% confidence interval [1.33-5.18],p=0.01) or life-threatening ventricular arrhythmias (12/22[55%] vs 27/124[21.8%], HR=3.10 [1.57-6.12],p=0.001) within 30 days after presentation. Compared to ACR, patients with ICI-myocarditis were more likely to experience life-threatening ventricular arrhythmias (22/147 [16.3%] vs 1/50 [2%];p=0.01) or third-degree heart block (25/147 [17.0%] vs 0/50 [0%];p=0.002). In ICI-myocarditis, overall mortality, myocarditis-related mortality, and composite outcome adjusted for age and sex were associated with pathological Q-waves on presenting ECG (hazard ratio by subdistribution model [HR(sh)]=5.98[2.8-12.79],p<.001; 3.40[1.38-8.33],p=0.008; 2.20[0.95-5.12],p=0.07; respectively) but inversely associated with Sokolow-Lyon Index (HR(sh)/mV=0.57[0.34-0.94],p=0.03; HR(sh)=0.54[0.30-0.97],p=0.04; 0.50[0.30-0.85],p=0.01; respectively). The composite outcome was also associated with conduction disorders on presenting ECG (HR(sh)=3.27[1.29-8.34],p=0.01).

**Conclusions:** ICI-myocarditis has more life-threatening arrhythmias than ACR and manifests as decreased voltage, conduction disorders, and repolarization abnormalities. Ventricular tachycardias, complete heart block, low-voltage, and pathological Q-waves were associated with adverse outcomes.

**NCT:** NCT04294771

**Key Points:** *Question:* What are the electrocardiographic manifestations of immune checkpoint inhibitor (ICI)-associated myocarditis? How do they compare to acute cellular rejection (ACR), which is resembling pathophysiologically to ICI-myocarditis? Which electrocardiographic features are associated with adverse outcomes?

*Findings:* ICI-myocarditis results in more frequent ventricular arrhythmias and high-degree atrioventricular blocks compared to ACR. Prolonged QRS intervals, decreased voltage, conduction disorders, and pathological Q-waves are predictors of adverse outcomes in ICI-associated myocarditis.

*Meaning:* ICI-associated myocarditis is a highly arrhythmogenic cardiomyopathy. Ventricular arrhythmias, conduction disorders, low-voltage, and pathological Q-waves are associated with a poor prognosis.

## Introduction

Immune checkpoint inhibitors (ICI) have transformed oncology care with nearly 50% of cancer patients eligible for ICI treatment.^1^ ICI unleash cytotoxic T-cells to achieve anti-tumor effects but can also cause T-cell and macrophage mediated myocarditis.^2–4^ A subset of ICI recipients (0.3% to 1.1%) experience myocarditis, a rare immune related adverse event (IrAE) that can cause cardiogenic shock and fatal arrhythmias.^5,6^ The diagnosis of ICI-myocarditis remains challenging.^2,7^ Cardiac magnetic resonance imaging (cMRI) and endomyocardial biopsy (EMB) are often difficult to obtain due to patients’ critical condition. Furthermore, sensitivity of cMRI is estimated at 48% with EMB also resulting in false negatives.^8^ A multimodal approach incorporating biomarker, echocardiographic, and electrocardiographic (ECG) findings may represent a high yield strategy in diagnosing ICI-related myocarditis.^9^ However, ECG findings in ICI-myocarditis have yet to be systematically described and their prognostic significance has not yet been studied. We set out to describe presenting ECG and telemetry events in patients with ICI-myocarditis given that arrhythmogenic events are routinely and easily identified in presenting patients. We compared these findings to ECG from a cohort of heart transplant recipients diagnosed with acute cellular rejection (ACR). We hypothesized that ICI-myocarditis would mimic the low-voltage and QRS prolongation seen in ACR.^4,10,11^ This hypothesis was grounded in the many pathologic similarities between ACR and ICI-myocarditis, including lymphocytic infiltration, a similarity that has motivated the use of similar immunosuppressive treatment strategies for both conditions, including corticosteroids and anti-T cell directed therapies.^2–4,12–17^ Additionally, we hypothesized that presenting ECG features in ICI-myocarditis would predict death and life-threatening ventricular arrhythmias.

## Methods

### ICI-Myocarditis Selection

A retrospective multicenter registry spanning 49 institutions across 11 countries was used to collect 147 cases of ICI-myocarditis (Supplemental Table 1) as defined by European Society of Cardiology criteria for clinically suspected myocarditis with recent ICI exposure.^18^ External collaborating institutions were identified through cardio-oncology departments, via a website created to collect cases of ICI-myocarditis (www.cardioonc.org), and by contacting authors of published case reports (<u>Supplementary Data Methods 1</u>). Clinical data was collected and shared by participating collaborators via a HIPPA-compliant REDCap web-based platform (IRB: 181337; *NCT04294771*).^19,20^ All 147 cases were analyzed for presence of arrhythmias throughout hospitalization as reported by treating physicians. ECG on admission was independently examined for 125 cases where ECG was obtained within 3 days of admission (Supplemental Figure 1). When multiple presenting ECG were available, ECG closest to presentation and without complete heart block or supraventricular arrhythmias were preferentially selected. Baseline ECG was defined as the most recent ECG obtained before ICI exposure and was available for independent examination in 52 cases.

### ACR selection

Heart transplants at Vanderbilt University Medical Center complicated by grade 2R or 3R acute cellular rejection were selected in reverse chronological order and spanned 2013-2019.^21^ Cases of concomitant humoral rejection were excluded. ECG obtained less than 10 days after heart transplantation or more than 3 days from diagnostic EMB were excluded. Donor and recipient characteristics were collected via chart review and the Organ Procurement and Transplantation Network database.

### ECG Interpretation

Two blinded cardiologists (BO, JA) systematically quantified standard ECG intervals (PR, QRS, QTc, Sokoloff-Lyon Index) and evaluated for relevant qualitative features. ECG features were aggregated on basis of pathophysiological relatedness (Supplemental Table 2). Inter- and intra-observer variability was excellent (intra-class correlation>0.8) for PR, QRS, QTc and Sokoloff measurements (<u>Supplemental Data Methods 2</u>).

### Statistical Analysis

Paired t-test and McNemar’s test were used to compare features of presenting ECG to baseline ECG. Non-parametric Wilcoxon and Chi-squared test was used to compare ECG features in ICI-myocarditis to ACR. The primary outcome was myocarditis-related mortality in thirty days. The secondary outcomes were 1) a composite of either myocarditis-related death or life-threatening arrhythmia in thirty days (defined as sustained ventricular tachycardia, ventricular fibrillation, torsade de pointes, pulseless electrical activity, or asystole) and 2) all-cause mortality in thirty days.

The primary outcome analysis used features on the presenting ECG as the independent variable. Since our methodology preferentially selects for ECG that do not exclusively capture heart block, life-threatening ventricular arrhythmias, or supraventricular arrhythmias, a focused secondary analysis used the aggregate incidence of these arrhythmias throughout the entire hospitalization as the independent variable to test association with outcomes of interest. In both analyses, Cox proportional-hazards model determined association with all-cause mortality over the 30-day surveillance period. Competing risk analysis (Subdistribution hazards model, i.e., Fine-Gray model) was used to account for mortality due to causes other than myocarditis for the outcomes of myocarditis-related mortality or composite outcome. These models were separately adjusted for age and sex in a multivariable analysis. Hazard Ratio (HR), 95% confidence interval, and cumulative incidence curves were presented.

## Results

### Demographics

The 147 patients with ICI-myocarditis had a median (IQR) age of 67 years (58-77) and 92/147 (62.6%) were male (Table 1). Median days from first ICI dose to myocarditis presentation was 38 days (21-83). In 146 patients with 30-day surveillance, 39/146 (26.7%) died within 30 days of presentation of which 24/39 (62%) of deaths were attributable to myocarditis. Other leading causes of death included to cancer progression - 6/39 (15%), sepsis - 6/39 (15%), and non-cardiac IrAE 7/39 (18%), of which 6/7 (86%) were attributable to non-cardiac myotoxicities (e.g., myositis). Pacemakers and/or defibrillators were placed in 22/146 (15.1%) patients within 30 days of presentation.

**Table 1.** ICI-myocarditis cases characteristics and outcomes.

|  | Total |
| --- | --- |
|  | Med (IQR) N; n/N (%) |
| Age | 67 (58-77)<br>N=147 |
| Female | 55/147 (37.4%) |
| Body Mass Index | 25.3 (21.4-28.8)<br>N=138 |
| Hyperlipidemia | 49/138 (35.5%) |
| Diabetes | 25/138 (18.1%) |
| Hypertension | 77/140 (55.0%) |
| Prior Tobacco User | 69/137 (50.4%) |
| Pre-existing Stroke | 5/138 (3.6%) |
| Pre-existing Peripheral Vascular Disease | 11/137 (8.0%) |
| Pre-existing Coronary Artery Disease | 27/139 (19.4%) |
| Pre-existing Heart Failure | 16/138 (11.6%) |
| 1 or More Traditional Cardiovascular Risk Factors (defined as HLD or DM2 or HTN or Tobacco use) | 115/140 (82.1%) |
| Prior History of Cardiac Disease (defined as CAD or CHF) | 34/137 (24.8%) |
| Prior History of Cardiovascular Disease (PVD, CVA, CAD, CHF or HTN) | 89/138 (64.5%) |
| Index ICI Therapy Category |  |
| - Anti CTLA-4 & PD1/PDL1 Combination Therapy | 27/147 (18.4%) |
| - Anti CTLA-4 Monotherapy | 41/147 (27.9%) |
| - Anti PD1/PDL1 Monotherapy | 79/147 (53.7%) |
| Days from First ICI Dose to Hospital Admission | 38 (21-83)<br>N=139 |
| Days from Last ICI Dose to Hospital Admission | 15 (9-22)<br>N=139 |
| Number of Doses ICI Received | 2 (1-4) N=140 |
| Cancer Type |  |
| - Bladder Cancer | 4/147 (2.7%) |
| - Breast Cancer | 1/147 (0.7%) |
| - Kidney Cancer | 16/147 (10.9%) |
| - Leukemia | 2/147 (1.4%) |
| - Lung Cancer | 52/147 (35.4%) |
| - Non-Hodgkin Lymphoma | 1/147 (0.7%) |
| - Prostate Cancer | 2/147 (1.4%) |
| - Melanoma | 40/147 (27.2%) |
| - Thymic Cancer (Non-Thymoma) | 2/147 (1.4%) |
| - Esophageal Cancer | 4/147 (2.7%) |
| - Gastric Cancer | 2/147 (1.4%) |
| - Colorectal Cancer | 1/147 (0.7%) |
| - Endometrial Cancer | 1/147 (0.7%) |
| - Hepatocellular Carcinoma | 2/147 (1.4%) |
| - Cholangiocarcinoma | 1/147 (0.7%) |
| - Squamous Cell Carcinoma | 4/147 (2.7%) |
| - Other Cancer | 1/147 (0.7%) |
| - Mesothelioma | 3/147 (2.0%) |
| - Thymoma | 8/147 (5.4%) |
| At Least One Other Concomitant IrAE | 102/147 (69.4%) |
| Concomitant IrAE: Myasthenia Gravis-Like Syndrome | 32/147 (21.8%) |
| Concomitant IrAE: Immune-Related Myositis / Rhabdomyolysis | 45/147 (30.6%) |
| Abnormal ECG<sup>18</sup> | 135/147 (91.8%) |
| Abnormal Troponin | 123/132 (93.2%) |
| Initial Troponin >10x Upper Limit of Normal | 81/126 (64.3%) |
| Reduced LVEF On Initial TTE Admission (LVEF<50%) | 59/141 (41.8%) |
| Reduced LVEF During Hospitalization For ICI-Myocarditis (LVEF<50%) | 66/141 (46.8%) |
| Cardiac Magnetic Resonance Imaging Compatible with Myocarditis | 54/75 (72%) |
| Cardiac Biopsy Proven Myocarditis | 29/40 (73%) |
| ***Cumulative Incidence of Arrhythmia Throughout Hospital Stay*** |  |
| Supraventricular Arrhythmia* | 35/147 (23.8%) |
| - Atrial Fibrillation | 31/147 (21.1%) |
| - Atrial Flutter | 2/147 (1.4%) |
| - Multifocal Atrial Tachycardia | 2/147 (1.4%) |
| Conduction Disorder* | 101/147 (68.7%) |
| - Bundle Branch or Fascicular Blocks | 90/147 (61.2%) |
| - First-Degree Heart Block | 23/147 (15.6%) |
| - Second-Degree Heart Block | 11/147 (7.5%) |
| - Third-Degree Heart Block | 25/147 (17.0%) |
| ECG Finding of Pericarditis (PR Depression or Diffuse ST Elevations) | 20/147 (13.6%) |
| Repolarization Abnormalities (ST-Segment Or T-Wave Changes) | 72/147 (49.0%) |
| Premature Ventricular Complexes (Any Type) | 41/147 (27.9%) |
| Ventricular Arrhythmias (Any Type; Sustained or Non-Sustained) | 25/147 (17.0%) |
| Life-Threatening Ventricular Arrhythmias* | 22/147 (15.0%) |
| - Asystole | 4/147 (2.7%) |
| - Pulseless Electrical Activity | 4/147 (2.7%) |
| - Ventricular Fibrillation | 4/147 (2.7%) |
| - Ventricular Tachycardia Unspecified Morphology, Sustained | 7/147 (4.8%) |
| - Ventricular Tachycardia Monomorphic, Sustained | 12/147 (8.2%) |
| - Ventricular Tachycardia Polymorphic, Sustained | 1/147 (0.7%) |

\* this category includes rhythms below and that patients may experience more than one of these rhythms

|  |  |
| --- | --- |
| - Ventricular Tachycardia Torsade De Pointes, Sustained | 2/147 (1.4%) |
| Third-Degree Heart Block and/or Life-Threatening Ventricular Arrhythmia | 36/147 (24.5%) |
| Third-Degree Heart Block and Life-Threatening Ventricular Arrhythmia | 11/147 (7.5%) |
| **Outcome** |  |
| Placement of a Pacemaker and/or Defibrillator Within 30 days | 22/146 (15.1%) |
| Pacemaker Without Defibrillator Within 30 days | 21/146 (14.4%) |
| Length of Stay (In Days) | 11 (7-24) N=98 |
| In-Hospital Mortality | 42/147 (28.6%) |
| 30-Day All-Cause Mortality | 39/146 (26.7%) |
| 30-Day Myocarditis-Related Mortality or Life-Threatening Ventricular Arrhythmia | 37/146 (25.3%) |
| Diagnostic Certainty<sup>9</sup> |  |
| - Definite Myocarditis | 81/143 (56.6%) |
| - Probable Myocarditis | 27/143 (18.9%) |
| - Possible Myocarditis | 35/143 (24.5%) |
| **Cause of Death<sup>†</sup> (Of 39 Patients With 30d All-Cause Mortality)** |  |
| Myocarditis | 24/39 (61.5%) |
| Cancer Progression | 6/39 (15.4%) |
| Immune Related Adverse Event Other Than Cardiotoxicity<sup>†</sup> | 7/39 (17.9%) |
| - Non-Cardiac Myotoxicities Including Myasthenia Gravis-Like Syndrome Associated with Diaphragmatic Failure | 6/7 (85.7%) |
| - Thrombocytopenia, Immune Related | 1/7 (14.3%) |
| Sepsis | 6/39 (15.4%) |
| Thromboembolic Event | 2/39 (5.1%) |
| Hemorrhage | 3/39 (7.7%) |
| Respiratory Failure (Other Than Diaphragmatic Failure)<sup>‡</sup> | 3/39 (7.7%) |
| - Pulmonary Infection | 2/3 (66.7%) |
| - Acute Respiratory Distress Syndrome | 2/3 (66.7%) |
| Ischemic Stroke | 1/39 (2.6%) |
| Unknown | 1/39 (2.6%) |

Abbreviations: CAD: coronary artery diseases; CHF: congestive heart failure; CTLA-4: Cytotoxic T-lymphocyte-associated protein 4; CVA: Cerebrovascular accident; HTN: Hypertension; ICI: Immune checkpoint inhibitor; IrAE: Immune Related Adverse Event; LVEF: Left ventricular ejection fraction; PDI: Programmed cell death protein 1; PD-L1: Programmed death-ligand 1; PVD: Peripheral vascular disease; TTE: Transthoracic echocardiogram
<sup>†</sup> note more than one cause may contribute to death
<sup>‡</sup> note more than one cause may contribute to respiratory failure

In total, 135/147 (91.8%) patients experienced abnormal ECG during hospitalization. Throughout hospitalization (median: 11 days, IQR:7-24), 101/147 (68.7%) patients experienced conduction disorders, which included second-degree heart block (11/147 (7.5%)) and complete heart block (25/147 (17.0%)). Of note, supraventricular arrhythmias had a cumulative incidence of 35/147 (23.8%). A total of 22/147 (15.0%) patients experienced life-threatening ventricular arrhythmia, including 16/147 (10.9%) sustained ventricular tachycardia, 4/147 (2.7%) ventricular fibrillation, 2/147 (1.4%) torsade de pointes, 4/147 (2.7%) pulseless electrical activity, and 4/147 (2.7%) asystole. A total of 11/147 (7.5%) patients developed both complete heart block and a life-threatening ventricular arrhythmia.

### Comparison to Baseline ECG

Baseline ECG obtained before ICI exposure was available for comparison in 52 cases. Paired analysis comparing presenting ECG to baseline ECG showed ICI-myocarditis presents with elevated heart rate (93.9 vs 80.4 bpm;p=0.009) and prolongation of QRS (95.3 vs 93.2 ms;p=0.02) and QT interval corrected for heart rate using Fridericia’s formula (441.8 vs 421.0 ms;p=0.03) (Table 2). There was a significant decrease in cardiac depolarization voltage assessed by the quantitative Sokolow-Lyonn Index (1.39 vs 1.69 mV;p=0.006). The incidence of left bundle branch block (LBBB) (10/52 [19%] vs 3/52 [6%];p=0.046) and sinus tachycardia (25/52 [48%] vs 15/52 [29%];p=0.02) were increased from baseline. In aggregate, conduction disorders (35/52 [67%] vs 23/52 [44%];p=0.01) and repolarization abnormalities (27/52 [52%] vs 13/52 [25%],p=0.008) were significantly increased. Of note, ECG suggestive of pericarditis were infrequent without significant increase from baseline (4/52 [8%] vs 1/52 [2%],p=0.25).

**Table 2:** Presenting ECG of ICI-myocarditis as compared to baseline and as predictors of myocarditis-related mortality and composite outcome using survival analyses adjusting for age and sex*.

|  | ICI-Myocarditis,<br>Presenting ECG | ICI-Myocarditis,<br>Baseline ECG |  | Subdistribution Hazards<br>Model For 30d Myocarditis-Related Mortality Adjusting<br>for Age and Sex | Subdistribution Hazards<br>Model For 30d Composite<br>Outcome Adjusting for Age<br>and Sex |
| --- | --- | --- | --- | --- | --- |
|  | Med (IQR) N;<br>n/N (%) | Med (IQR) N;<br>n/N (%) | p-value<br>(paired T-test) | HR(sh) [95% CI], p-value* | HR(sh) [95% CI], p-value* |
| Heart Rate (bpm) | 93.9 [72.6-114.7]<br>N=52 | 80.4 [68.1-94.8]<br>N=52 | 0.009 | 1.01 [0.99-1.03], p=.52 N=125 | 1.00 [0.99-1.02], p=.60 N=125 |
| PR Length (ms) | 162.8 [136.0-186.0] N=42 | 154.1 [136.0-187.6] N=46 | 0.10 | 1.00 [0.99-1.02], p=.90 N=107 | 1 [0.99-1.01], p=.62 N=107 |
| QTcF Length (ms) | 441.8 [414.9-462.6] N=49 | 421.0 [399.2-440.4] N=51 | 0.03 | 1.00 [0.99-1.01], p=.59 N=122 | 1.00 [1.00-1.01], p=.42 N=122 |
| QRS Length (ms) | 95.3 [85.7-118.2]<br>N=52 | 93.2 [82.7-102.5]<br>N=52 | 0.02 | 1.01 [0.99-1.02], p=.57 N=125 | 1.01 [1-1.03], p=.03 N=125 |
| Sokolow-Lyon Index (mV) | 1.39 [0.85-2.03]<br>N=52 | 1.69 [1.28-2.26]<br>N=52 | 0.006 | 0.54 [0.30-0.97], p=.04 N=124 | 0.50 [0.30-0.85], p=.01 N=124 |
|  |  |  | p-value<br>(McNemar's<br>test) |  |  |
| **CONDUCTION DISORDERS<sup>†</sup>** | 35/52 (67%) | 23/52 (44%) | 0.01 | 1.91 [0.71-5.14], p=.20 N=125 | 3.27 [1.29-8.34], p=.01 N=125 |
| - Bundle Branch Block, Left<br>Bundle | 10/52 (19%) | 3/52 (6%) | 0.05 | 0.85 [0.26-2.79], p=.79 N=125 | 1.49 [0.62-3.61], p=.37 N=125 |
| - Bundle Branch Block, Right<br>Bundle | 14/52 (27%) | 9/52 (17%) | 0.18 | 1.63 [0.69-3.85], p=.27 N=125 | 2.22 [1.06-4.67], p=.04 N=125 |
| - Fascicular Block, Left<br>Anterior | 10/52 (19%) | 5/52 (10%) | 0.23 | 1.58 [0.57-4.41], p=.38 N=125 | 1.81 [0.82-3.97], p=.14 N=125 |
| - Fascicular Block, Left<br>Posterior | 6/52 (12%) | 2/52 (4%) | 0.22 | 1.40 [0.47-4.14], p=.54 N=125 | 1.56 [0.52-4.62], p=.43 N=125 |

\* Only arrhythmia subgroups with at least n>2 in ICI-myocarditis presenting ECG are shown
† When multiple eligible ECG were available, ECG without complete heart block or supraventricular arrhythmias were preferentially selected for this analysis focusing on PR, QRS and QTc measurements. Please see Table 1 for cumulative incidence of arrhythmias in ICI-myocarditis.

|  |  |  |  |  |  |
| --- | --- | --- | --- | --- | --- |
| - Heart Block, First Degree | 9/52 (17%) | 7/52 (13%) | 0.72 | 1.78 [0.57-5.58], p=.32 N=125 | 2.14 [0.83-5.53], p=.12 N=125 |
| **ECG Findings of Pericarditis** | 4/52 (8%) | 1/52 (2%) | 0.25 | 0.58 [0.14-2.40], p=.46 N=125 | 0.98 [0.34-2.82], p=.97 N=125 |
| - ST Segment Elevation, Diffuse | 3/52 (6%) | 1/52 (2%) | 0.62 | 0.63 [0.15-2.61], p=.52 N=125 | 1.05 [0.36-3.05], p=.93 N=125 |
| **PREMATURE VENTRICULAR COMPLEX (ALL TYPES)** | 9/52 (17%) | 3/52 (6%) | 0.08 | 1.36 [0.43-4.32], p=.61 N=125 | 1.95 [0.74-5.10], p=.18 N=125 |
| - Premature Ventricular Complex | 9/52 (17%) | 3/52 (6%) | 0.08 | 0.96 [0.27-3.38], p=.95 N=125 | 1.51 [0.56-4.07], p=.42 N=125 |
| **SINUS MECHANISM** | 42/52 (81%) | 46/52 (88%) | 0.29 | 0.58 [0.21-1.59], p=.29 N=125 | 0.70 [0.29-1.70], p=.43 N=125 |
| - Normal Sinus Rhythm | 17/52 (33%) | 31/52 (60%) | 0.002 | 0.43 [0.16-1.16], p=.09 N=125 | 0.61 [0.28-1.32], p=.21 N=125 |
| - Sinus Tachycardia | 25/52 (48%) | 15/52 (29%) | 0.02 | 1.48 [0.6-3.65], p=.39 N=125 | 1.28 [0.61-2.68], p=.52 N=125 |
| **REPOLARIZATION ABNORMALITIES** | 27/52 (52%) | 13/52 (25%) | 0.008 | 1.57 [0.64-3.89], p=.33 N=125 | 1.48 [0.68-3.24], p=.33 N=125 |
| - ST Segment Depression, Diffuse | 5/52 (10%) | 1/52 (2%) | 0.22 | 0.66 [0.09-4.73], p=.68 N=125 | 0.47 [0.07-3.27], p=.44 N=125 |
| - ST Segment Depression, Regional | 4/52 (8%) | 0/52 (0%) | NA | 1.04 [0.13-8.56], p=.97 N=125 | 1.48 [0.35-6.32], p=.59 N=125 |
| - T Wave Inversions | 21/52 (40%) | 12/52 (23%) | 0.07 | 1.98 [0.81-4.82], p=.13 N=125 | 1.42 [0.63-3.24], p=.40 N=125 |
| **SUPRAVENTRICULAR ARRHYTHMIA<sup>†</sup>** | 7/52 (13%) | 6/52 (12%) | 1.00 | 2.84 [0.99-8.16], p=.052 N=125 | 2.39 [1.01-5.65], p=.047 N=125 |
| - Atrial Fibrillation<sup>†</sup> | 6/52 (12%) | 5/52 (10%) | 1.00 | 2.19 [0.67-7.24], p=.20 N=125 | 2.11 [0.77-5.76], p=.14 N=125 |
| **UNCATEGORIZED** |  |  |  |  |  |
| Premature Atrial Complex | 5/52 (10%) | 3/52 (6%) | 0.68 | 2.19 [0.57-8.45], p=.26 N=125 | 1.63 [0.49-5.43], p=.42 N=125 |
| Left Ventricular Hypertrophy | 12/52 (23%) | 16/52 (31%) | 0.34 | 0.71 [0.21-2.43], p=.58 N=125 | 0.51 [0.16-1.63], p=.25 N=125 |
| Low QRS Voltage | 4/52 (8%) | 1/52 (2%) | 0.37 | 6.05 [2.10-17.39], p<.001 N=125 | 2.70 [0.97-7.49], p=.06 N=125 |
| P Wave Abnormality Suggestive of Left Atrial Enlargement | 11/52 (21%) | 9/52 (17%) | 0.75 | 1.40 [0.53-3.71], p=.49 N=125 | 1.09 [0.46-2.59], p=.85 N=125 |
| Q Waves, Pathological | 8/52 (15%) | 4/52 (8%) | 0.22 | 3.40 [1.38-8.33], p=.008 N=125 | 2.20 [0.95-5.12], p=.07 N=125 |

### Outcome Analysis by Cumulative Incidence of Arrhythmia

Patients with ICI-myocarditis were more likely to experience all-cause mortality within 30 days if they developed complete heart block (12/25 [48%] vs 27/122 [22.1%]; HR=2.62, 95% confidence interval=[1.33-5.18],p=0.01) or life-threatening ventricular arrhythmias (12/22 [55%] vs 27/125 [21.6%]; HR=3.10 [1.57-6.12],p=0.001) at any point during hospitalization (cumulative incidence curves in Figure 1).

**Figure 1.**
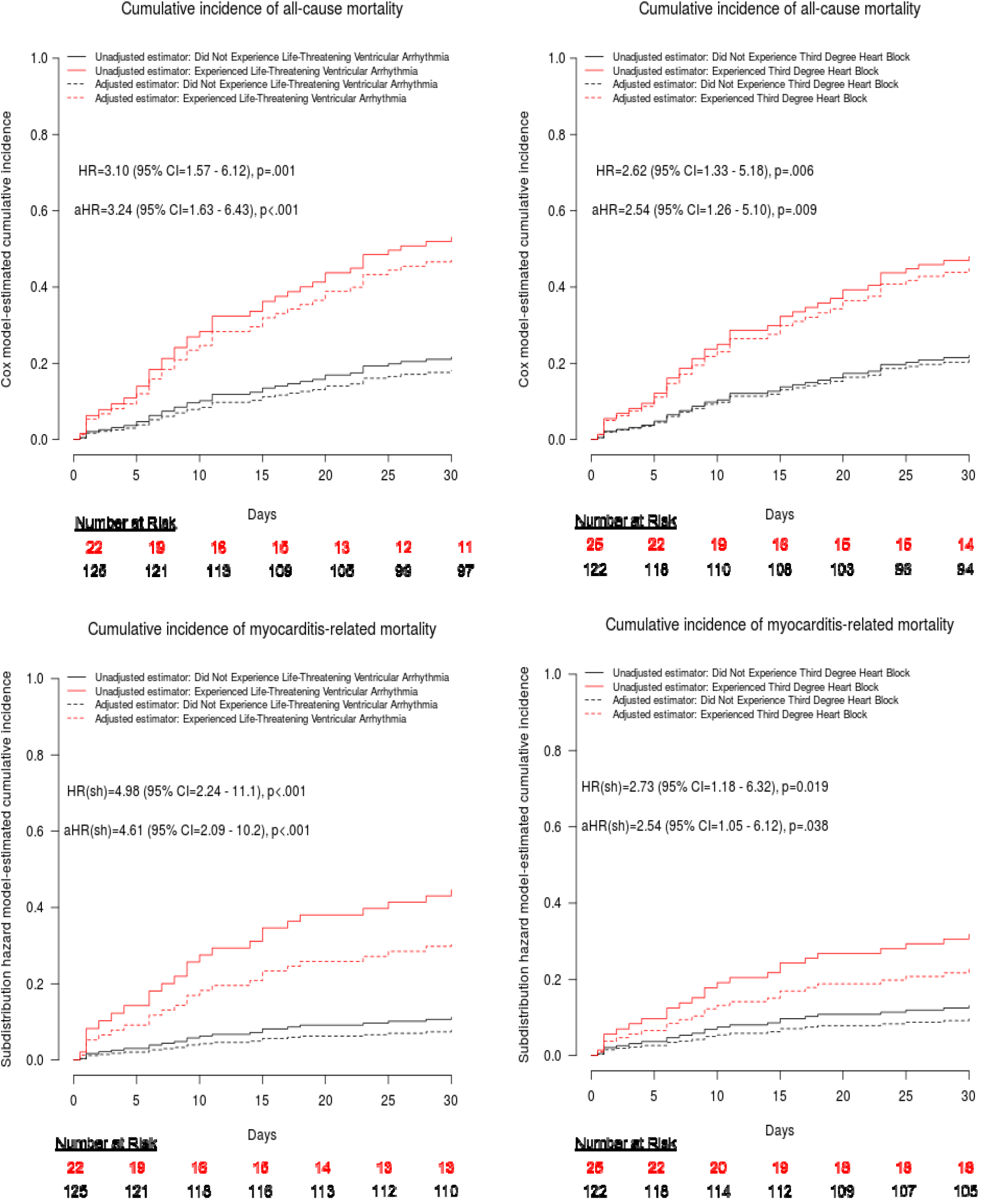
Outcomes by cumulative incidence of arrhythmia.

Additionally, myocarditis-related mortality within 30 days was more common in patients who developed complete heart block (8/25 [32%] vs 16/122 [13.1%]; hazard ratio by subdistribution model[HR(sh)=2.73 [1.18-6.32],p=0.019) or life-threatening ventricular arrhythmias (10/22 [45.5%] vs 14/125 [11.2%]; HR(sh)=4.98 [2.24-11.1],p<0.001) (cumulative incidence curves in Figure 1).

Composite outcome of myocarditis-related mortality or life-threatening ventricular arrhythmia within 30 days was also more common in patients who experienced complete heart block (13/25 [52%] vs 24/122 [19.7%]; HR(sh)=3.55 [1.80-6.99],p<0.001) (figure not shown).

Supraventricular arrhythmia at any point during hospitalization was not associated with either all-cause mortality (13/35 [37%] vs 26/112 [23.2%]; HR=1.67 [0.86-3.25],p=0.13), myocarditis-related mortality (8/35 [22.9%] vs 16/112 [14.3%]; HR(sh)=1.61 [0.71-3.7],p=0.26), or composite outcome within 30 days (13/35 [37.1%] vs 24/112 [21.4%]; HR(sh)=1.72 [0.91-3.26],p=0.10) (cumulative incidence curves in Supplemental Figure 2).

### Outcome Analysis by Presenting ECG Features

A total of 125 ICI-myocarditis patients met criteria to be included in the analysis of predictive value of presenting ECG features and 22 were excluded due to initial ECG obtained more than 3 days from admission or initial ECG with paced rhythm or exclusively capturing ventricular tachycardia (flow chart of analyzed ECG in Supplemental Figure 1, characteristics of the population in Supplemental Table 3). Using survival analyses, thirty-day myocarditis-related mortality was significantly associated with pathological Q-waves (7/19 [37%] vs 13/106 [12.3%]; HR(sh)=3.67 [1.46-9.22],p=0.006) and low QRS voltage (3/6 [50%] vs 17/119 [14.3%]; HR(sh)= 4.50 [1.34-15.12],p=0.02) and showed a trend towards inverse association with Sokolow-Lyon Index (HR(sh)/mV=0.55 [0.28-1.06],p=0.08) (cumulative incidence curves in Figure 2, model results in Supplemental Table 4, cumulative incidence curves by Sokolow-Lyon Index in Supplemental Figure 3).

**Figure 2.**
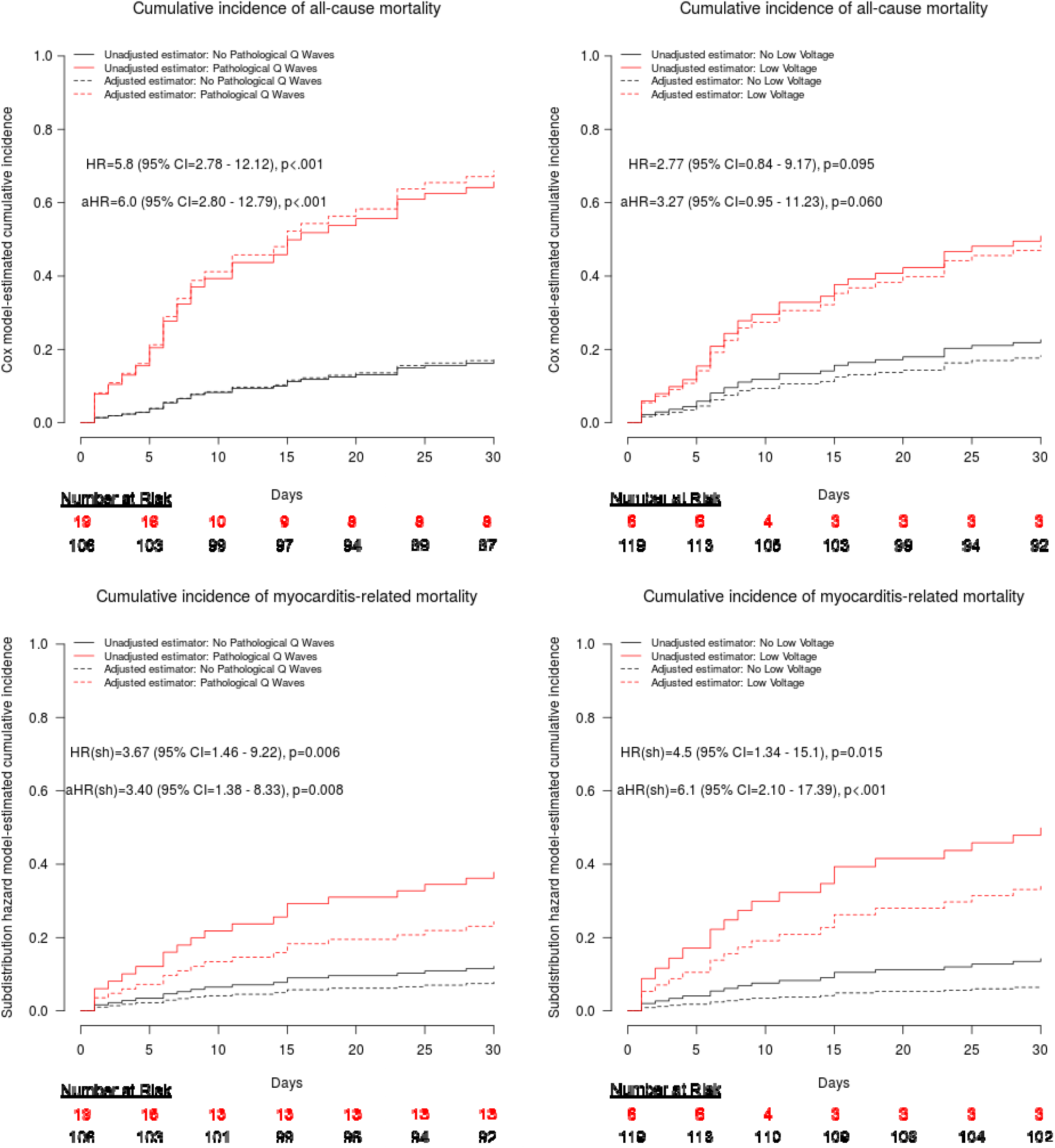
Outcomes by presenting ECG findings.

Using survival analyses, composite outcome of myocarditis-related mortality or life-threatening ventricular arrhythmia was inversely associated with Sokolow-Lyon Index (HR(sh)/mV=0.51 [0.30-0.87],p=0.01) and positively associated with RBBB (14/43 [33%] vs 14/82 [17%]; HR(sh)=2.16 [1.05-4.47],p=0.04) and conduction disorders generally (23/79 [29%] vs 5/46 [11%]; HR(sh)=3.05 [1.20-7.76],p=0.02) (cumulative incidence curves in Supplemental Figure 4, model results in Supplemental Table 4, cumulative incidence curvesby Sokolow-Lyon Index in Supplemental Figure 3). Composite outcome of myocarditis-related mortality or life-threatening ventricular arrhythmia showed a trend towards association with pathological Q-waves (7/19 [37%] vs 21/106 [19.8%]; HR(sh)=2.10 [0.90-4.89],p=0.09) and low QRS voltage (3/6 [50%] vs 25/119 [21.0%]; HR(sh)= 2.57 [0.90-7.28],p=0.08).

Similarly, all-cause mortality was associated with pathological Q-waves (12/19 [63%] vs 18/106 [17.0%]; HR=5.80 [2.78-12.12],p<0.001) and inversely associated with Sokolow-Lyon Index (HR/mV=0.59 [0.35-0.98],p=0.04) (cumulative incidence curves in Figure 2, model results in Supplemental Table 4, cumulative incidence curves by Sokolow-Lyon Index in Supplemental Figure 3).

Multivariable survival analysis was performed by adding covariates of age and sex into cox proportional-hazards model and sub distribution hazards models. This analysis mirrored the results of survival analyses described above (myocarditis-related mortality & composite outcome: Table 2, all-cause mortality: Supplemental Table 5; Figures 1 & 2; Supplemental Figures 2 & 3).

### Comparison to ACR

The 50 patients with ACR had median (IQR) age of 51 years (43-62), 64% (32/50) of whom were male (Supplemental Table 6). Median days from transplant to ACR was 145 days (IQR:26-283). 29/50 (58%) were admitted during or as a result of ACR, with median length of stay of 12 days (IQR:5-21). 2R rejection was seen in 46/50 (92%) and 4/50 (8%) had 3R rejection. Throughout hospitalization (if applicable) or at presenting ECG, 34/50 (68%) patients experienced conduction disorders but second or third-degree heart block was not seen in any patients. There was a cumulative incidence of 6/50 (12%) supraventricular arrhythmias and 1/50 (2%) life-threatening ventricular arrhythmia. None of the patients required a pacemaker and/or defibrillator within 30 days after ACR diagnosis.

Compared to ACR, ECG at the time of ICI-myocarditis had comparable voltage and QRS duration (Table 3). ICI-myocarditis had significantly more LBBB (20/125 [16.0%] vs 0/50 [0%];p=0.003) and left anterior fascicular block (LAFB) (24/125 [19.2%] versus 3/50 (6%];p=0.02) but fewer right bundle branch block (RBBB) (43/125 [34.4%] vs 27/50 [54%];p=0.02), and right atrial abnormality (4/125 [3.2%] vs 10/50 [20%];p<.001). In aggregate, ICI-myocarditis had more premature ventricular contractions (PVCs) (18/125 [14.4%] vs 1/50 [2%];p=0.02) but fewer repolarization abnormalities (53/125 [42.4%] vs 33/50 [66%];p=0.005). ACR was less severe than ICI-myocarditis in terms of 30-day all-cause mortality (0/50 [0%] vs 39/146 [26.7%];p<0.001), in-hospital incidence of left ventricular ejection fraction less than 50% (4/28 [14.3%] vs 66/141 [46.8%];p=0.001), progression to severe life-threatening ventricular arrhythmias at admission or during hospital stay (1/50 [2%] vs 22/147 [16.3%];p=0.01), and pacemaker or defibrillator placement within 30 days of the ACR or ICI-myocarditis event (0/50 [0%] vs 22/146 [11.1%];p=0.004). Additionally, ACR had a lower cumulative incidence of third-degree heart block (0/50 [0%] vs 25/147 [17.0%];p=0.002) compared to ICI-myocarditis.

**Table 3:** Comparison on ECG findings in ICI-myocarditis to acute cellular rejection at presentation.

|  | **ICI-Myocarditis, Presenting ECG**<br>**N=125** | **Acute Cellular Rejection 2R/3R Presenting ECG**<br>**N=50** |  |
| --- | --- | --- | --- |
|  |  |  | p-value (Wilcoxon test) |
| Heart Rate (bpm) | 87.6 [71.3-104.6] N=125 | 88.8 [80.4-110.2] N=50 | 0.20 |
| PR Interval Length (ms) | 161.3 [145.7-180.6] N=107 | 153.2 [136.5-166.1] N=48 | **0.01** |
| QTcF Length (ms) | 432.5 [405.4-462.1] N=122 | 434.1 [393.5-460.1] N=49 | 0.59 |
| QRS Length (ms) | 95.0 [85.3-122.3] N=125 | 92.8 [85.5-103.2] N=49 | 0.15 |
| Sokolow-Lyon Index | 1.240 [0.700-1.889] N=124 | 1.421 [0.889-1.845] N=50 | 0.40 |
|  |  |  | p-value (Chi-square test) |
| **CONDUCTION DISORDERS** | 79/125 (63%) N=125 | 34/50 (68%) N=50 | 0.55 |
| - Bundle Branch Block, Left Bundle | 20/125 (16%) N=125 | 0/50 (0%) N=50 | **0.003** |
| - Bundle Branch Block, Nonspecific | 2/125 (2%) N=125 | 2/50 (4%) N=50 | 0.34 |
| - Bundle Branch Block, Right Bundle | 43/125 (34%) N=125 | 27/50 (54%) N=50 | **0.02** |
| - Escape Rhythm, Ventricular | 1/125 (1%) N=125 | 0/50 (0%) N=50 | 0.53 |
| - Fascicular Block, Left Anterior | 24/125 (19%) N=125 | 3/50 (6%) N=50 | **0.03** |
| - Fascicular Block, Left Posterior | 13/125 (10%) N=125 | 4/50 (8%) N=50 | 0.63 |
| - Heart Block, First Degree | 18/125 (14%) N=125 | 5/50 (10%) N=50 | 0.44 |
| - Heart Block, Third Degree* | 5/125 (4%) N=125 | 0/50 (0%) N=50 | 0.15 |

\* When multiple eligible ECG were available, ECG without complete heart block or supraventricular arrhythmias were preferentially selected for this analysis focusing on PR, QRS and QTc measurements. Please see Table 1 for cumulative incidence of arrhythmias in ICI-myocarditis and Supplemental-Table-3 for cumulative incidence of arrhythmias in ACR.

|  |  |  |  |
| --- | --- | --- | --- |
| **ECG FINDINGS OF PERICARDITIS** | 17/125 (14%) N=125 | 2/50 (4%) N=50 | 0.07 |
| - PR-Segment Depression | 1/125 (1%) N=125 | 0/50 (0%) N=50 | 0.53 |
| - ST Segment Elevation, Diffuse | 16/125 (13%) N=125 | 2/50 (4%) N=50 | 0.08 |
| **PREMATURE VENTRICULAR COMPLEX (ALL TYPES)** | 18/125 (14%) N=125 | 1/50 (2%) N=50 | **0.02** |
| - Premature Ventricular Complex | 17/125 (14%) N=125 | 1/50 (2%) N=50 | **0.02** |
| - Premature Ventricular Complex Bigeminy | 2/125 (2%) N=125 | 0/50 (0%) N=50 | 0.37 |
| **SINUS MECHANISM** | 107/125 (85.6%) N=125 | 47/50 (94%) N=50 | 0.08 |
| - Sinus Tachycardia | 51/125 (40.8%) N=125 | 21/50 (42%) N=50 | 0.81 |
| **REPOLARIZATION ABNORMALITIES** | 53/125 (42%) N=125 | 33/50 (66%) N=50 | **0.005** |
| - ST Segment Elevation, Regional | 8/125 (6%) N=125 | 0/50 (0%) N=50 | 0.07 |
| - ST Segment Depression, Diffuse | 9/125 (7%) N=125 | 2/50 (4%) N=50 | 0.43 |
| - ST Segment Depression, Regional | 7/125 (6%) N=125 | 3/50 (6%) N=50 | 0.92 |
| - T Wave Inversions | 41/125 (33%) N=125 | 29/50 (58%) N=50 | **0.002** |
| - T Wave Notching | 0/125 (0%) N=125 | 1/50 (2%) N=50 | 0.11 |
| **SUPRAVENTRICULAR ARRHYTHMIA**Error! Bookmark not defined. | 11/125 (9%) N=125 | 2/50 (4%) N=50 | 0.27 |
| - Atrial FibrillationError! Bookmark not defined. | 10/125 (8%) N=125 | 1/50 (2%) N=50 | 0.14 |
| - Atrial Flutter**Error! Bookmark not defined.** | 1/125 (1%) N=125 | 1/50 (2%) N=50 | 0.50 |
| **UNCATEGORIZED** |  |  |  |
| Premature Atrial Complex | 8/125 (6%) N=125 | 0/50 (0%) N=50 | 0.07 |
| Premature Junctional Complex | 1/125 (1%) N=125 | 0/50 (0%) N=50 | 0.53 |
| Left Ventricular Hypertrophy | 21/125 (17%) N=125 | 10/50 (20%) N=50 | 0.62 |
| Low QRS Voltage | 6/125 (5%) N=125 | 2/50 (4%) N=50 | 0.82 |
| P Wave Abnormality Suggestive of Left Atrial Enlargement | 29/125 (23%) N=125 | 14/50 (28%) N=50 | 0.51 |
| P Wave Abnormality Suggestive of Right Atrial Enlargement | 4/125 (3%) N=125 | 10/50 (20%) N=50 | **<0.001** |
| Q-waves, Pathological | 19/125 (15%) N=125 | 4/50 (8%) N=50 | 0.20 |
| Accelerated Junctional Rhythm | 1/125 (1%) N=125 | 0/50 (0%) N=50 | 0.53 |

## Discussion

In this study, we assessed ECG features of ICI-myocarditis using a large international database. We show that ICI-myocarditis manifests as clinically significant electrocardiographic disturbances including high degree heart block and ventricular arrhythmias, which are strongly associated with poor clinical outcomes. Compared to baseline ECG, there are also other ECG manifestations, including repolarization abnormalities, decreased voltage, and increases in heart rate, QRS, and QTc. Low-voltage, conduction disorders, and pathological Q-waves were predictive of myocarditis-related death, life-threatening cardiac arrhythmias, and/or overall mortality.

This is the first study to systematically analyze ECG in ICI-myocarditis from a large number of patients with ICI-associated myocarditis with two cardiologists systematically quantifying and evaluated the ECG while blinded to the clinical features for each patient.

Previous cohort studies had reported electrical disturbances as a major clinical feature of ICI-associated myocarditis.^6,8,22^ Our finding that 91.8% of patients have abnormal ECG is supported by Mahmoud et al’s cohort of 35 patients where 89% of patients had abnormal ECG.^6^ In addition, our finding that 42% of patients present with ST-segment or T wave abnormalities was similar to the 37% in Escudier et al.’s 30 patient cohort and 55% in Zhang et. al’s 103 patient cohort.^8,23^ In addition, Zhang et al found 80% of patients presented in sinus rhythm with a cumulative incidence of complete heart block of 16% compared to 86% and 17% respectively in our cohort.^8^

Although we hypothesized that the electrophysiological manifestations of ICI-myocarditis would resemble those of ACR, given the striking pathological similarities, our results show that ICI-myocarditis is both more arrhythmogenic and more lethal than ACR. Life-threatening ventricular arrhythmias, PVCs, and conduction disorders affecting the left ventricle including complete heart block were more common in ICI-myocarditis but not a major feature of ACR.

Interestingly, our study also represents the largest description of ECG findings in moderate-severe ACR. While previous studies have correlated ACR with atrial arrhythmias, sustained ventricular arrhythmias, PR, QRS, and QT lengthening, these changes were infrequently seen in presenting ECG among our cohort.^22,24^ Instead, most ECG changes could be explained by post-surgical changes, including sinus tachycardia, P-wave enlargement, right bundle branch block, and nonspecific ST changes.^24^ While low voltage and pathological Q waves were infrequent, they were not significantly different from the ICI-myocarditis cohort, suggesting that both immune infiltrates had similar electromotive effects despite differing impact on electrical conduction.

Our prognostic analysis adds to and is supportive of predictive ECG studies in general myocarditis. While several studies of myocarditis due to heterogenous causes have shown pathological Q-waves to be predictive of fulminant myocarditis, they did not find significant association with long-term survival.^25,26^ While studies have shown that low-voltage lacks predictive value for death in allograft rejection, it has not previously been studied in myocarditis.^10,27^ It is interesting that while Rassi et. al found Chagas heart disease to have a 9% prevalence of low-voltage with a hazard ratio for mortality of 1.87, we found a similar prevalence of 8% in ICI-myocarditis but with much higher hazard ratio for mortality of approximatively 4.5.^28^ This may be explained by differences in acuity between these two inflammatory cardiomyopathies as well as the relatively denser inflammatory infiltrates in ICI-myocarditis.^2,29^

Both low-voltage and pathological Q-waves signify a loss of electromotive force and are intuitive markers for the extent of inflammatory infiltrate and cardiomyocyte damage. Unlike low-voltage where there is a global decrease in electrical current, Q-waves represent potentials from the unaffected ventricular wall opposite to an inflammatory focus that has become electrically inert. The finding that these two features are strong predictors of mortality suggests that suppressing the underlying inflammatory infiltrate may be a greater priority than antiarrhythmic drugs or devices.

ICI-myocarditis is histologically characterized by dense, patchy infiltrates of lymphocytes and macrophages that affect both the myocardium and the conduction system.^2^ Compared with ACR, which is primarily lymphocytic, ICI-myocarditis is characterized by both lymphocyte and macrophage infiltrates with a higher CD68/CD3 (macrophages/lymphocytes) ratio.^3^ Denser infiltrates in ICI-myocarditis are associated with increased myocyte necrosis and a different molecular profile with lower macrophage expression of PD-L1 perhaps reflecting an influx of the reparative M2 macrophage subpopulation.^3^ Importantly, macrophages have been shown to electrically couple with cardiomyocytes even in the absence of disease, thereby facilitating depolarization and improving AV conduction.^30^ It is possible that changes in macrophage phenotype and density in ICI-myocarditis may mediate the high frequency of conduction system blocks and ventricular ectopy seen in our cohort. Mouse models of ICI-myocarditis have replicated arrhythmogenicity and lympho-histiocytic infiltration seen in humans and may offer future insights into the electrical contribution of immune cells in inflammatory cardiomyopathies.^31^ Separately, other novel forms of cancer immunotherapy also demonstrate high levels of arrhythmogenicity; ventricular tachycardias and atrial fibrillation are disproportionately reported in CAR-T therapy while 20% of patients receiving IL-2 therapy developed arrhythmias requiring pharmacological intervention.^32–35^ These examples further illustrate how the emerging relationship between the immune system and cardiac conduction will become increasingly important in treatment of patients receiving immunotherapy and as a target for arrhythmia management more broadly.

Although this study would not have been possible without a multicenter approach, this introduced variability in data collection and interpretation. To mitigate this effect, clear criteria for adjudication were provided and each submission was subjected to a bi-institutional review process. Self-reporting allowed us to assemble an ICI-myocarditis cohort of this size but likely selected for more clinically severe cases. To account for this in our comparison to ACR, we excluded Grade 1R rejection. Nevertheless, our findings are less generalizable to low-severity cases of ICI-myocarditis. The comparison to baseline ECG was limited by availability of baseline ECG which likely enriched for patients with pre-existing cardiac disease thereby underestimating ECG changes caused by ICI. Our analysis only interprets initial ECG and thus does fully capture the predictive value of ECG changes that develop during hospitalization.

Although we were unable to correct for variance in treatment in the outcome analysis, we believe that the composite outcome of life-threatening ventricular arrhythmia or myocarditis-related death helps mitigate this by capturing early events that would have led to death if not for aggressive therapy.

## Conclusions

On ECG, ICI-myocarditis manifests as diffuse alteration of the cardiac conduction system represented by conduction blocks, decrease in QRS voltage, and appearance of cardiomyocyte death with pathological Q-waves. These features predict severe life-threatening ventricular arrhythmias and death. Clinicians should focus on identifying these ECG changes as part of multimodal diagnostic workup for ICI-myocarditis. Patients with these features are at higher risk for adverse outcomes and may benefit from more aggressive treatment and monitoring strategies.

## Data Availability

The datasets generated during and/or analysed during the current study are available from the corresponding author on reasonable request.

## Acknowledgments

We would like to thank all the collaborators who have participated in this multicenter database (Supplemental Table 1). This study was supported by the following grants: UL1 TR000445 from NCATS/NIH.

## Supplemental Data

**Supplemental Table 1.**
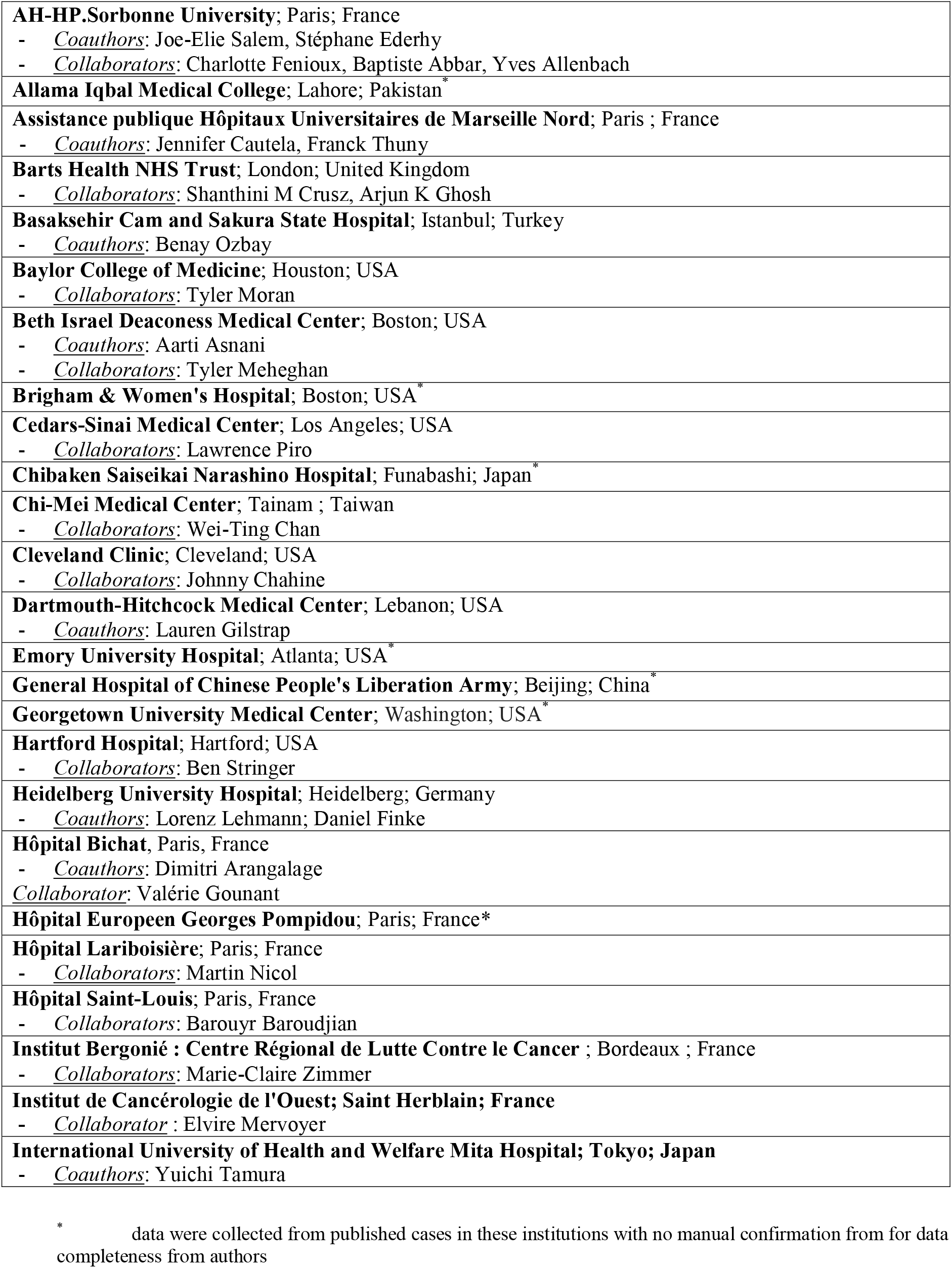
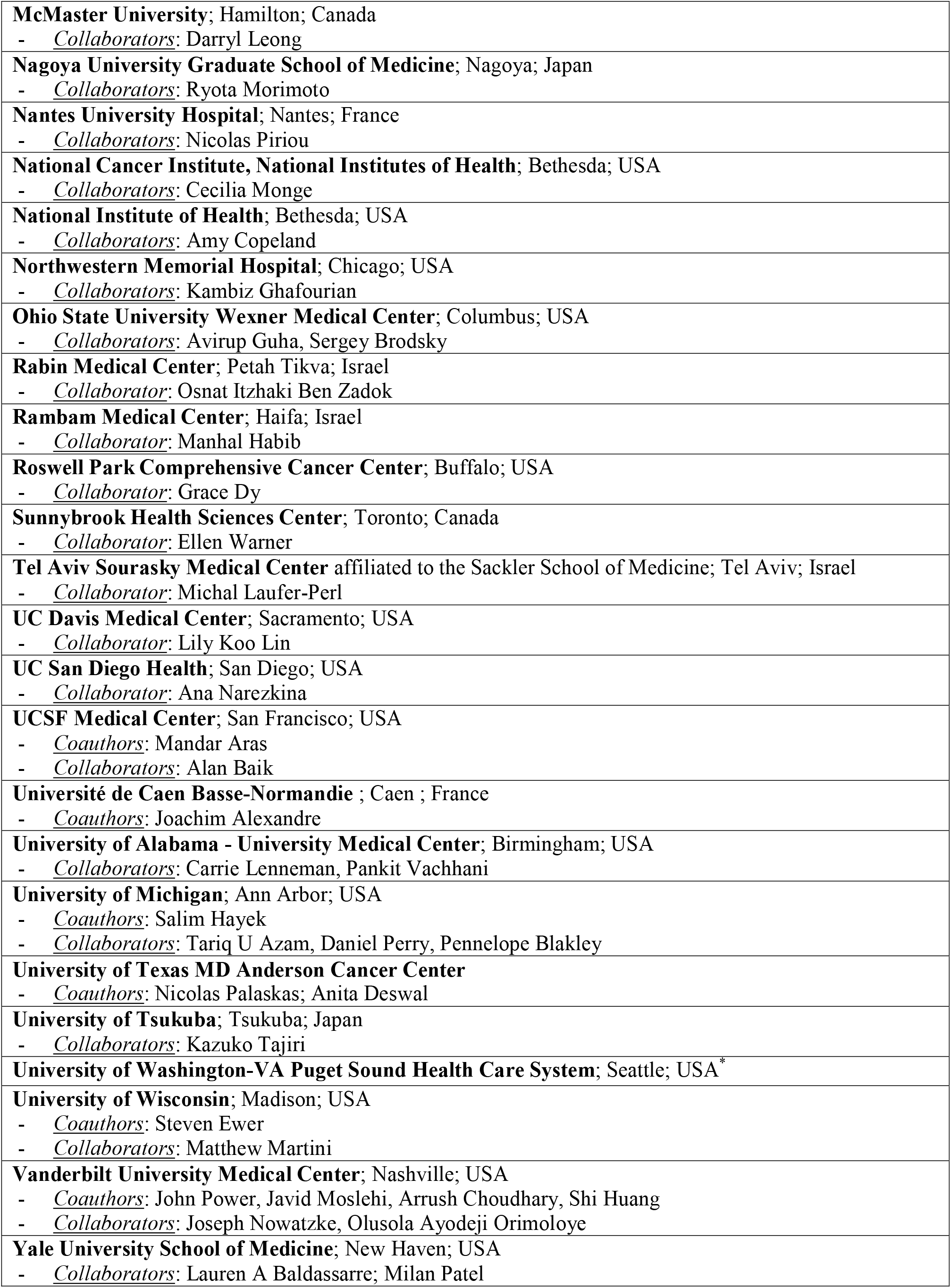
List of participating institutions.

**Supplemental Table 2:**
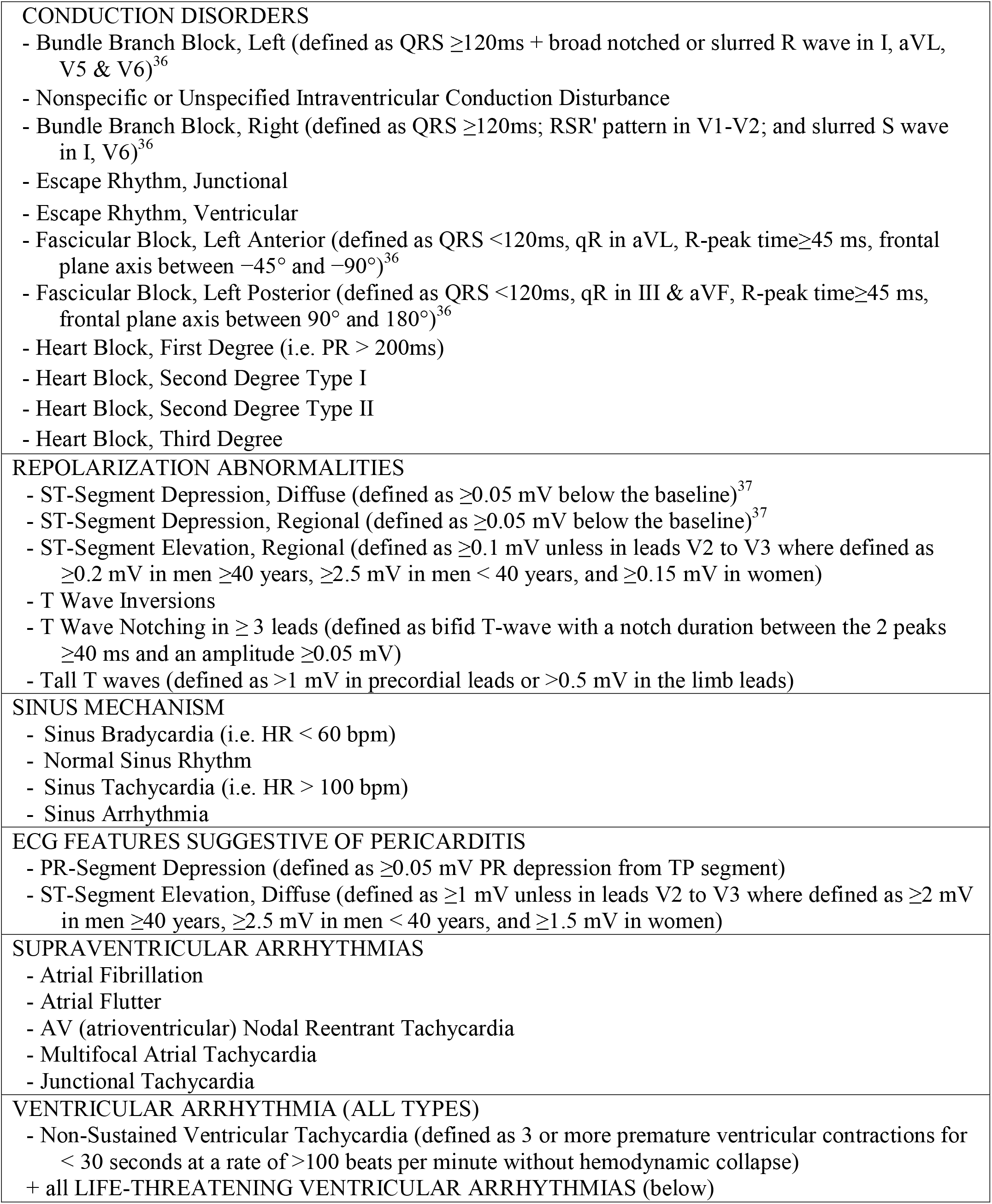
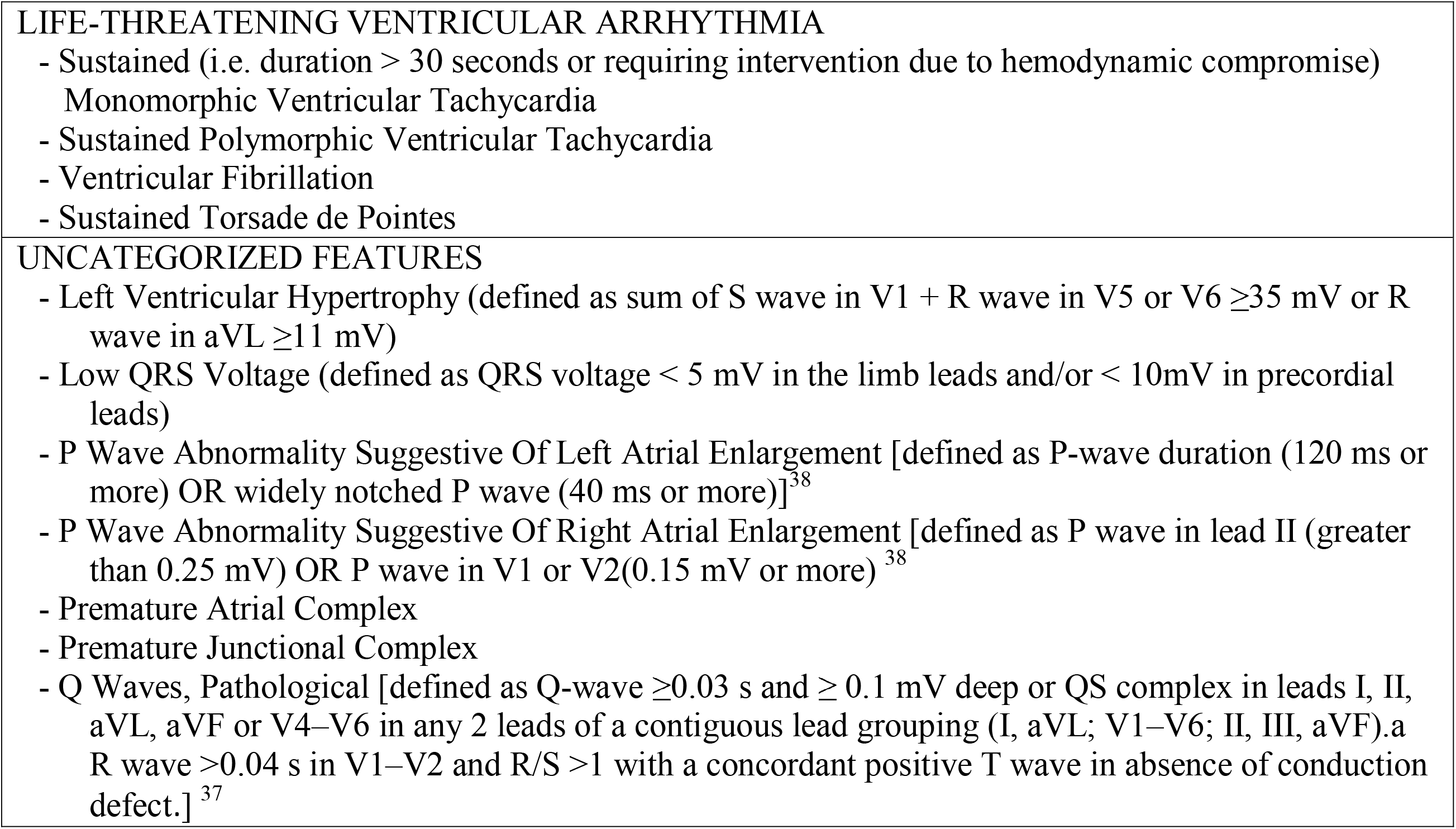
Glossary of qualitative ECG findings by category.

**Supplemental Table 3:** Cumulative incidence of arrhythmia throughout hospital stay for 125 ICI-myocarditis patients in ECG features quantitative outcome analysis. (Please refer to Supplemental Table 2 for full details on diagnostic criteria and categorization of qualitative ECG features)

|  | n/N (%) |
| --- | --- |
| **SINUS MECHANISM** | 107/125 (85.6%) |
| - Normal Sinus Rhythm | 56/125 (44.8%) |
| - Sinus Bradycardia | 2/125 (1.6%) |
| - Sinus Tachycardia | 55/125 (44.0%) |
| - Sinus Arrhythmia | 1/125 (0.8%) |
| **CONDUCTION DISORDERS** | 87/125 (69.6%) |
| - Bundle Branch Block, Nonspecific | 8/125 (6.4%) |
| - Bundle Branch Block, Left Bundle | 23/125 (18.4%) |
| - Bundle Branch Block, Right Bundle | 45/125 (36.0%) |
| - Escape Rhythm, Ventricular | 4/125 (3.2%) |
| - Escape Rhythm, Junctional | 4/125 (3.2%) |
| - Fascicular Block, Left Anterior | 25/125 (20.0%) |
| - Fascicular Block, Left Posterior | 14/125 (11.2%) |
| - Heart Block, First Degree | 19/125 (15.2%) |
| - Heart Block, Second Degree Type I | 4/125 (3.2%) |
| - Heart Block, Second Degree Type II | 5/125 (4.0%) |
| - Heart Block, Third Degree | 19/125 (15.2%) |
| **ECG FINDINGS OF PERICARDITIS** | 18/125 (14.4%) |
| - PR-Segment Depression | 1/125 (0.8%) |
| - ST Segment Elevation, Diffuse | 17/125 (13.6%) |
| **REPOLARIZATION ABNORMALITIES** | 62/125 (49.6%) |
| - ST segment elevation, regional | 13/125 (10.4%) |
| - ST Segment Depression, Diffuse | 11/125 (8.8%) |
| - ST Segment Depression, Regional | 8/125 (6.4%) |
| - Tall T Waves | 1/125 (0.8%) |
| - T Wave Inversions | 45/125 (36.0%) |
| - T Wave Notching | 5/125 (4.0%) |
| **VENTRICULAR EXCITABILITY (PVC or Ventricular Arrhythmia)** | 42/125 (33.6%) |
| **PREMATURE VENTRICULAR COMPLEX (ALL TYPES)** | 33/125 (26.4%) |
| - Premature Ventricular Complex | 31/125 (24.8%) |
| - Premature Ventricular Complex Bigeminy | 5/125 (4.0%) |
| - Premature Ventricular Complex Trigeminy | 1/125 (0.8%) |
| **VENTRICULAR ARRHYTHMIAS (all types)** | 18/125 (14.4%) |
| - Ventricular Tachycardia, Non-Sustained | 9/125 (7.2%) |
| - Ventricular Tachycardia, Sustained | 9/125 (7.2%) |
| **LIFE-THREATENING VENTRICULAR ARRHYTHMIA** | 15/125 (12.0%) |
| - Asystole | 4/125 (3.2%) |
| - Pulseless Electrical Activity | 4/125 (3.2%) |
| - Ventricular Fibrillation | 4/125 (3.2%) |
| - Ventricular Tachycardia Unspecified Morphology, Sustained | 5/125 (4.0%) |
| - Ventricular Tachycardia Monomorphic, Sustained | 5/125 (4.0%) |
| - Ventricular Tachycardia Polymorphic, Sustained | 1/125 (0.8%) |
| - Ventricular Tachycardia Torsade de Pointes, Sustained | 2/125 (1.6%) |
| **SUPRA-VENTRICULAR ARRHYTHMIAS** | 30/125 (24.0%) |
| - Atrial Fibrillation | 26/125 (20.8%) |
| - Atrial Flutter | 2/125 (1.6%) |
| - Multifocal Atrial Tachycardia | 2/125 (1.6%) |
| - AV Nodal Reentrant Tachycardia | 2/125 (1.6%) |
| - Junctional Tachycardia | 0/125 (0.0%) |
| **UNCATEGORIZED** |  |
| Accelerated Idioventricular Rhythm | 3/125 (2.4%) |
| Accelerated Junctional Rhythm | 1/125 (0.8%) |
| Left Ventricular Hypertrophy | 22/125 (17.6%) |
| Low QRS Voltage | 12/125 (9.6%) |
| Q-Waves, Pathological | 22/125 (17.6%) |
| P Wave Abnormality Suggestive of Left Atrial Enlargement | 29/125 (23.2%) |
| P Wave Abnormality Suggestive of Right Atrial Enlargement | 4/125 (3.2%) |
| Premature Atrial Complex | 14/125 (11.2%) |
| Premature Junctional Complex | 2/125 (1.6%) |
| Sinus Arrest / Sinus Pause | 2/125 (1.6%) |
| Placement of a Pacemaker and/or Defibrillator Within 30 days | 19/124 (15.3%) |
| In-Hospital Mortality | 33/125 (26.4%) |
| 30-Day All-Cause Mortality | 30/124 (24.2%) |
| 30-Day Myocarditis-Related Mortality or Life-Threatening Ventricular Arrhythmia | 28/124 (22.6%) |
| **Cause of Death (of 30 patients with 30d all-cause mortality)** |  |
| Myocarditis | 20/30 (66.7%) |
| Cancer Progression | 6/30 (20%) |
| Immune Related Adverse Event Other Than Cardiotoxicity* | 6/30 (20%) |
| - Non-Cardiac Myotoxicities Including Myasthenia Gravis-Like Syndrome Associated with Diaphragmatic Failure | 5/6 (83%) |
| - Thrombocytopenia, Immune Related | 1/6 (17%) |
| Sepsis | 4/30 (13%) |
| Thromboembolic Event | 2/30 (7%) |
| Hemorrhage | 1/30 (3%) |
| Respiratory Failure (Other Than Diaphragmatic Failure)† | 2/30 (7%) |
| - Pulmonary Infection | 1/2 (50%) |
| - Acute Respiratory Distress Syndrome | 1/2 (50%) |
| Ischemic Stroke | 1/30 (3%) |
| Unknown | 1/30 (3%) |

\* note more than one cause may contribute to death
† note more than one cause may contribute to respiratory failure

**Supplemental Table 4:**
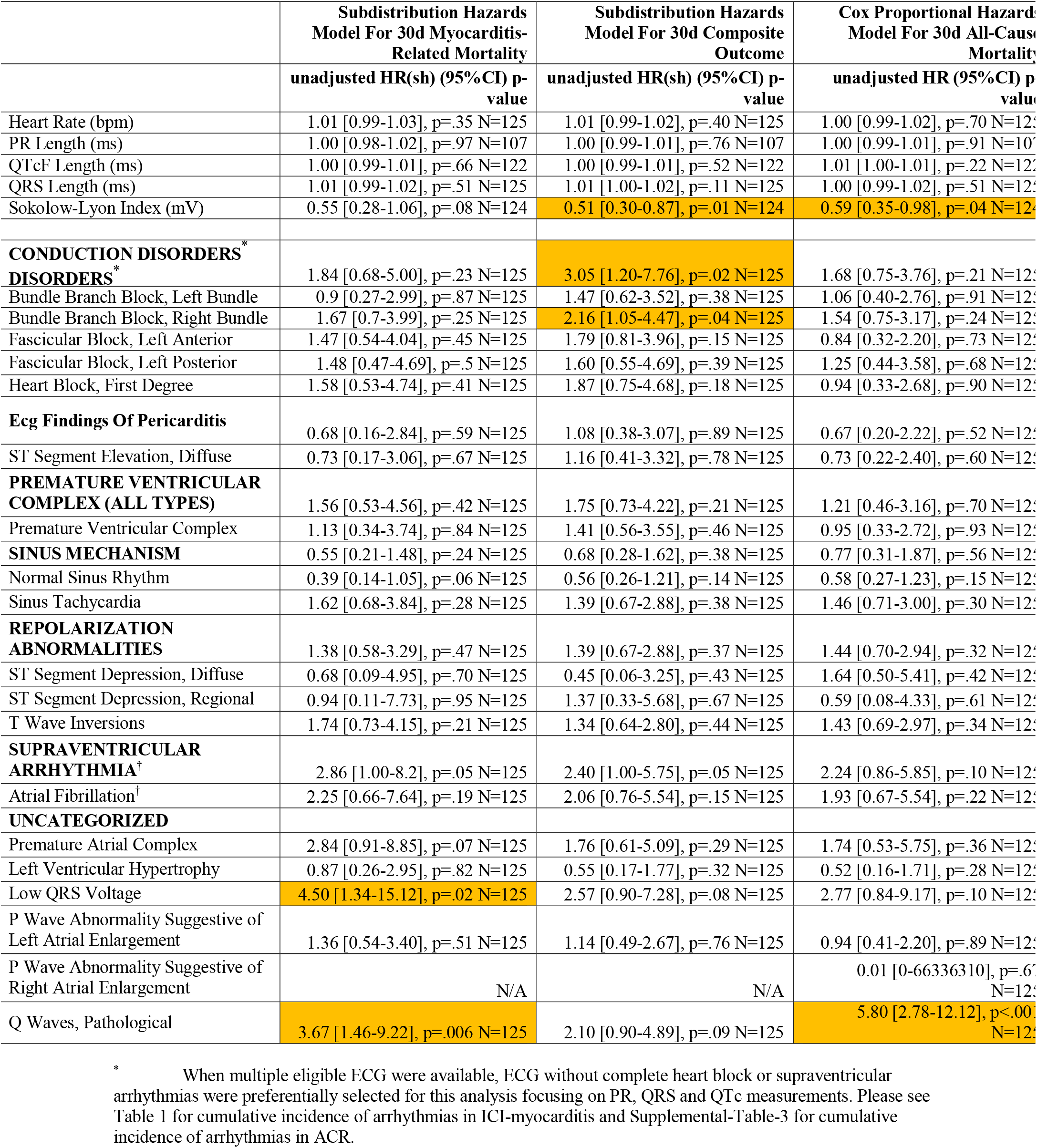
Presenting ECG of ICI-myocarditis as predictors of all-cause mortality, myocarditis-related mortality, and composite outcome using unadjusted survival analyses.

**Supplemental Table 5:** Presenting ECG of ICI-myocarditis as predictors of all-cause mortality using survival analyses adjusted for age and sex.

|  | **Cox Proportional Hazards**<br>**Model For 30d All-Cause**<br>**Mortality:**<br>**HR [95%CI], P-Value<sup>\*</sup>** |
| --- | --- |
| Heart Rate (bpm) | 1.01 [0.99-1.02], p=.40 N=125 |
| PR Length (ms) | 1.00 [0.99-1.01], p=.55 N=107 |
| QTcF Length (ms) | 1.00 [1.00-1.01], p=.36 N=122 |
| QRS Length (ms) | 1.00 [0.99-1.01], p=.90 N=125 |
| Sokolow-Lyon Index (mV) | **0.57 [0.34-0.94], p=.03 N=124** |
| **CONDUCTION DISORDERS<sup>†</sup>** | 1.56 [0.69-3.53], p=.29 N=125 |
| - Bundle Branch Block, Left Bundle | 1.00 [0.38-2.62], p=.99 N=125 |
| - Bundle Branch Block, Right Bundle | 1.48 [0.71-3.06], p=.29 N=125 |
| - Fascicular Block, Left Anterior | 0.85 [0.32-2.25], p=.75 N=125 |
| - Fascicular Block, Left Posterior | 1.34 [0.47-3.85], p=.59 N=125 |
| - Heart Block, First Degree | 0.83 [0.28-2.40], p=.72 N=125 |
| **ECG Findings Of Pericarditis** | 0.75 [0.22-2.51], p=.64 N=125 |
| - ST Segment Elevation, Diffuse | 0.83 [0.25-2.81], p=.76 N=125 |
| **PREMATURE VENTRICULAR COMPLEX (ALL TYPES)** | 1.01 [0.37-2.75], p=.99 N=125 |
| - Premature Ventricular Complex | 0.77 [0.26-2.30], p=.64 N=125 |
| **SINUS MECHANISM** | 0.77 [0.31-1.89], p=.56 N=125 |
| - Normal Sinus Rhythm | 0.50 [0.23-1.09], p=.08 N=125 |
| - Sinus Tachycardia | 1.67 [0.80-3.49], p=.17 N=125 |
| **REPOLARIZATION ABNORMALITIES** | 1.52 [0.74-3.12], p=.26 N=125 |
| ST Segment Depression, Diffuse | 1.60 [0.48-5.30], p=.44 N=125 |
| ST Segment Depression, Regional | 0.53 [0.07-3.90], p=.53 N=125 |
| T Wave Inversions | 1.49 [0.71-3.12], p=.29 N=125 |
| **SUPRAVENTRICULAR ARRHYTHMIA<sup>†</sup>** | 2.21 [0.84-5.79], p=.11 N=125 |
| - Atrial Fibrillation<sup>†</sup> | 1.83 [0.63-5.27], p=.27 N=125 |
| **UNCATEGORIZED** |  |
| Premature Atrial Complex | 1.59 [0.47-5.38], p=.46 N=125 |
| Left Ventricular Hypertrophy | 0.49 [0.15-1.61], p=.24 N=125 |
| Low QRS Voltage | 3.27 [0.95-11.23], p=.06 N=125 |
| P Wave Abnormality Suggestive of Left Atrial Enlargement | 1.10 [0.46-2.63], p=.83 N=125 |
| P Wave Abnormality Suggestive of Right Atrial Enlargement | 0.01 [0-77149830], p=.66 N=125 |
| Q Waves, Pathological | **5.98 [2.8-12.79], p<.001 N=125** |

\* Please see Table 1 for cumulative incidence of arrhythmias in ICI-myocarditis and Supplemental-Table-3 for cumulative incidence of arrhythmias in ACR.
<sup>†</sup> When multiple eligible ECG were available, ECG without complete heart block or supraventricular arrhythmias were preferentially selected for this analysis focusing on PR, QRS and QTc measurements. Please see Table 1 for cumulative incidence of arrhythmias in ICI-myocarditis and Supplemental-Table-3 for cumulative incidence of arrhythmias in ACR.

**Supplemental Table 6:** Baseline characteristics of acute cellular rejection cohort.

|  | Med (IQR) N; n/N (%) |
| --- | --- |
| Recipient Age, Years | 51 (43-62) N=50 |
| Female Recipient | 18/50 (36%) |
| Reason for Transplant |  |
| - Dilated Cardiomyopathy | 4/50 (8%) |
| - Ischemic Cardiomyopathy | 18/50 (36%) |
| - Amyloidosis | 1/50 (2%) |
| - Restrictive Cardiomyopathy | 1/50 (2%) |
| - Congenital Heart Disease | 4/50 (8%) |
| - Non-Ischemic Cardiomyopathy, Not Otherwise Specified | 17/50 (34%) |
| - Hypertrophic Cardiomyopathy | 2/50 (4%) |
| - Other | 3/50 (6%) |
| Donor Age | 29.0 (22.0-37.0) N=50 |
| Female Donor | 13/50 (26%) |
| Known Cardiac Allograft Vasculopathy | 11/50 (22%) |
| Induction Therapy |  |
| - Basiliximab (Simulect) | 26/50 (52%) |
| - Thymoglobulin (ATG) | 3/50 (6%) |
| - None | 20/50 (40%) |
| - Other | 1/50 (2%) |
| Background/Maintenance Immunosuppressive Regimen |  |
| - Prednisone + Tacrolimus + Mycophenolate | 42/50 (84%) |
| - Prednisone + Cyclosporine + Mycophenolate | 3/50 (6%) |
| - Other | 5/50 (10%) |
| Days from Transplant To Rejection | 145 (26-283) N=50 |
| Acute Cellular Rejection Grading Scheme<sup>21</sup> |  |
| - 2R, Moderate | 46/50 (92%) |
| - 3R, Severe | 4/50 (8%) |
| Days from Transplant To ECG | 145 (28-283) N=50 |
| Days from Biopsy To ECG | 0 (0-1) |
| 30-Day All-Cause Mortality | 0/50 (0%) |
| Placement of A Pacemaker and/or Defibrillator for ACR Related Arrhythmias Within 30 Days Of Diagnosis | 0/50 (0%) |
| Pacemaker Without Defibrillator for ACR Related Arrhythmias Within 30 Days Of Diagnosis | 0/50 (0%) |
| Admitted During or As A Result Of ACR | 29/50 (58.0%) |
| Length of Stay (Days)<sup>‡</sup> | 12 (5-21) N=29 |
| Reduced LVEF At Admission Or During Hospital Stay For ACR (Excluding Pre-Transplant LVEF)* | 4/28 (14.3%) |
| In-Hospital Mortality* | 0/29 (0%) |
| **Arrhythmias at Any Point During Hospitalization (If Applicable) Or At Presenting ECG (Please Refer To Supplemental Table 2 For Criteria / Classification)** |  |
| Supraventricular Arrhythmia<sup>§</sup> | 6/50 (12%) |
| - Atrial Fibrillation | 3/50 (6%) |

‡ This refers to the subset of admitted patients

|  |  |
| --- | --- |
| - Atrial Flutter | 2/50 (4%) |
| - Multifocal Atrial Tachycardia | 1/50 (2%) |
| Conduction Disorder<sup>§</sup> | 34/50 (68%) |
| - Bundle Branch or Fascicular Blocks | 33/50 (66%) |
| - First-Degree Heart Block | 6/50 (12%) |
| - Second-Degree Heart Block | 0/50 (0%) |
| - Third-Degree Heart Block | 0/50 (0%) |
| ECG Finding of Pericarditis (PR Depression Or Diffuse ST Elevations) | 2/50 (4%) |
| Repolarization Abnormalities (ST-Segment Or T-Wave Changes) | 33/50 (66%) |
| Premature Ventricular Complexes (Any Type) | 6/50 (12%) |
| Ventricular Arrhythmias (Any Type; Sustained or Non-Sustained) | 5/50 (10%) |
| Life-Threatening Ventricular Arrhythmias<sup>§</sup> | 1/50 (2%) |
| - Asystole | 0/50 (0%) |
| - Pulseless Electrical Activity | 0/50 (0%) |
| - Ventricular Fibrillation | 0/50 (0%) |
| - Ventricular Tachycardia Unspecified Morphology, Sustained | 0/50 (0%) |
| - Ventricular Tachycardia Monomorphic, Sustained | 1/50 (2%) |
| - Ventricular Tachycardia Polymorphic, Sustained | 0/50 (0%) |
| - Ventricular Tachycardia Torsade De Pointes, Sustained | 0/50 (0%) |
| Third-Degree Heart Block and/or Life-Threatening Ventricular Arrhythmia | 1/50 (2%) |

<sup>§</sup> this category includes rhythms below, note that patients may experience more than one of these rhythms

### Supplemental Data Methods 1: Systematic review search terms

Pubmed, Scopus, and Google Scholar were queried for case reports published between 1/1/2008 and 5/21/2019 with the search terms myocarditis, cardiotoxicity or cardiac toxicity in addition to (AND) at least one of the following: immune checkpoint inhibitor, pembrolizumab, ipilimumab, nivolumab, avelumab, atezolizumab, durvalumab, tremelimumab, anti-CTLA-4, anti-PD-L1, anti-PD-1, CTLA-4 inhibitor, PD-L1 inhibitor, OR PD-1 inhibitor.

### Supplemental Data Methods 2: ECG interval measurement

The QT interval was measured using the tangent method from the beginning of the QRS complex to the end of the T-wave. Lead II was preferentially used, but when unsuitable, V5 and V6 were used. The average of three consecutive PQRST complexes was used for each interval’ measurements. PVCs were excluded. In the rare cases in which three consecutive complexes were not available, two complexes were used. The heart rate corrected QT interval (QTc) was calculated using Bazett’s (QTcB=QT interval/) and Fredericia’s formula (QTcF=QT interval/(RR interval)^1/3^.

**Figure: ECG measurement with EP Calipers application (note that values used were an average of measurements across three consecutive PQRST complexes)**

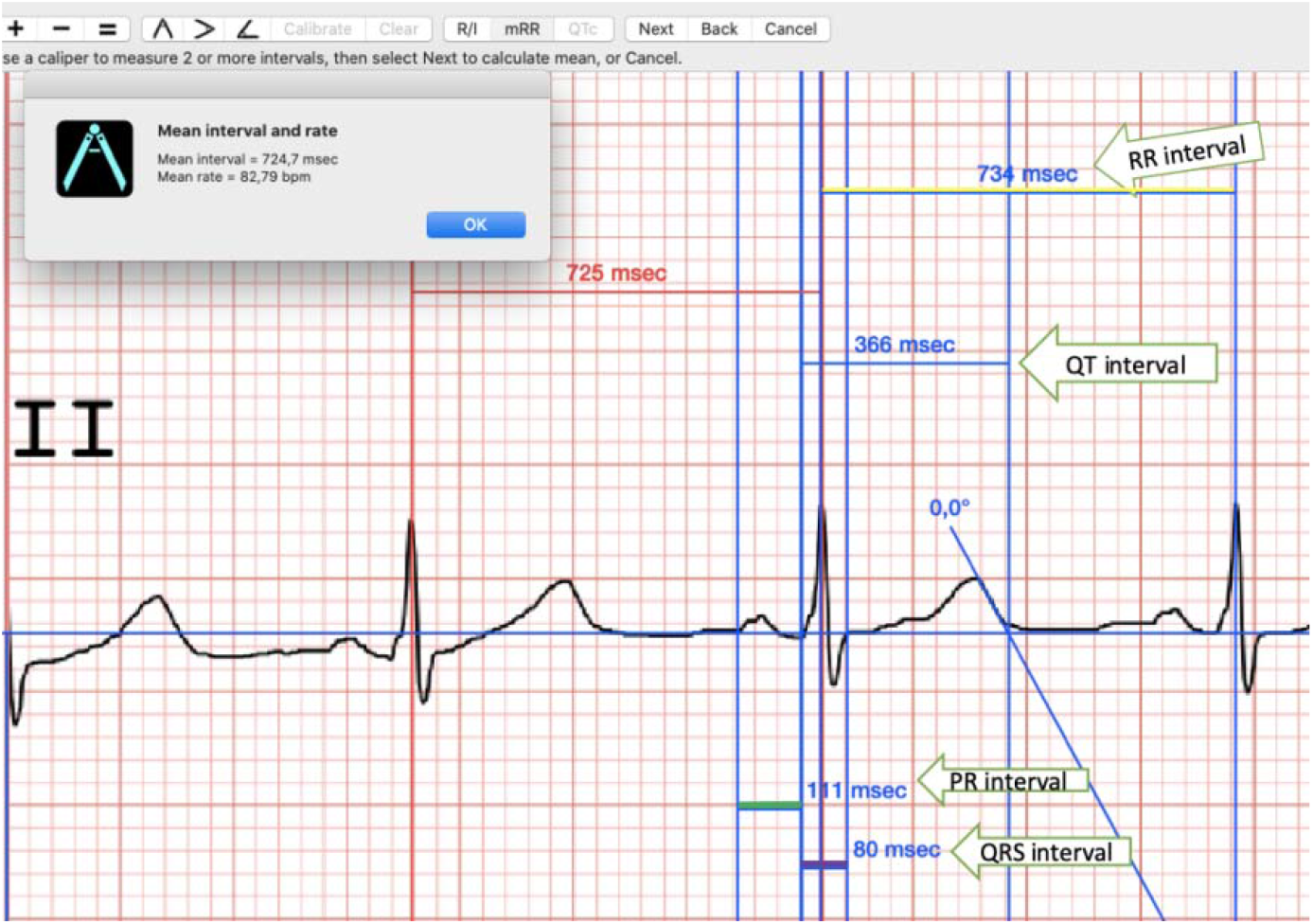

**Supplemental Figure 1:**
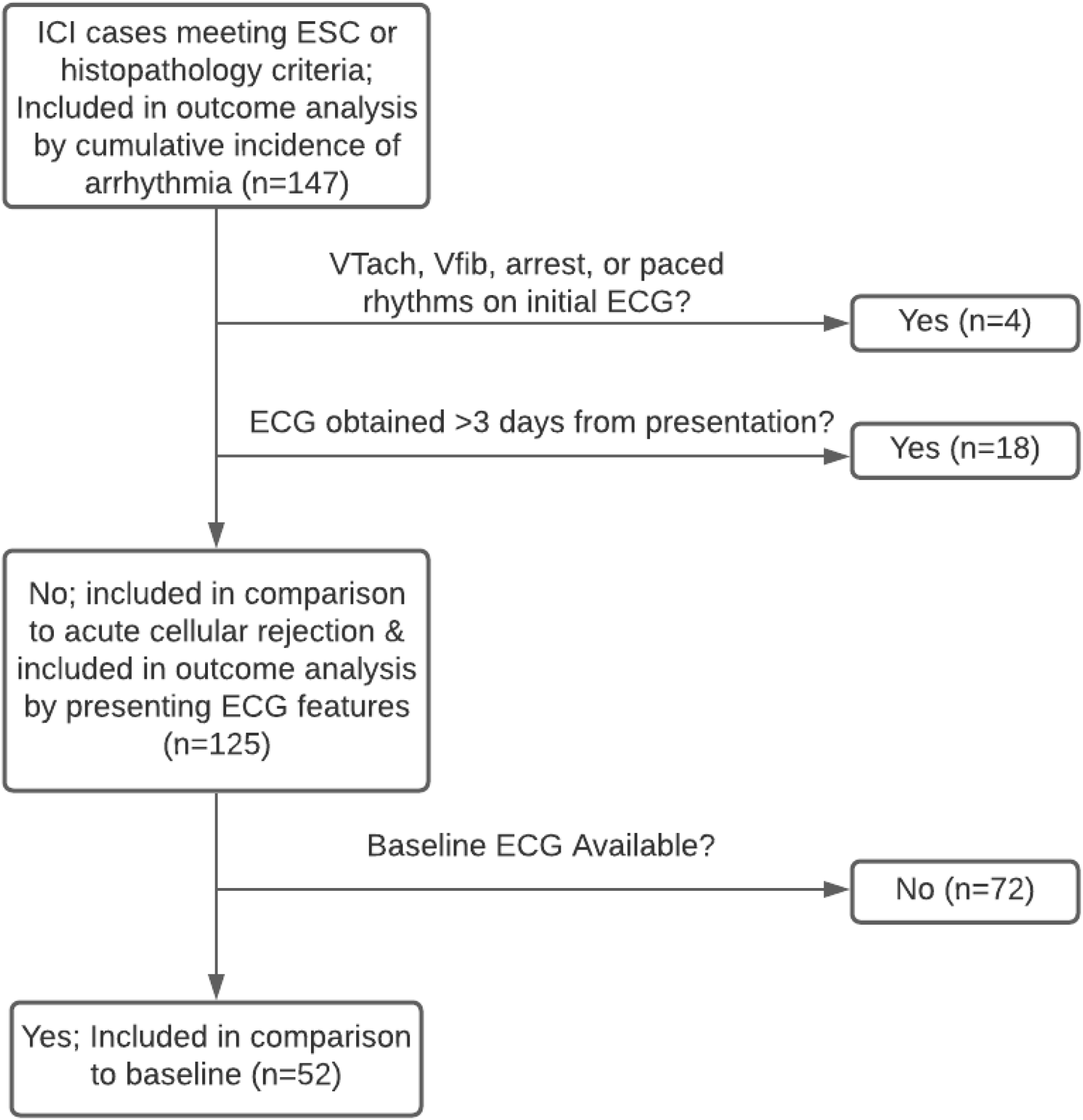
Flowchart.

**Supplemental Figure 2:**
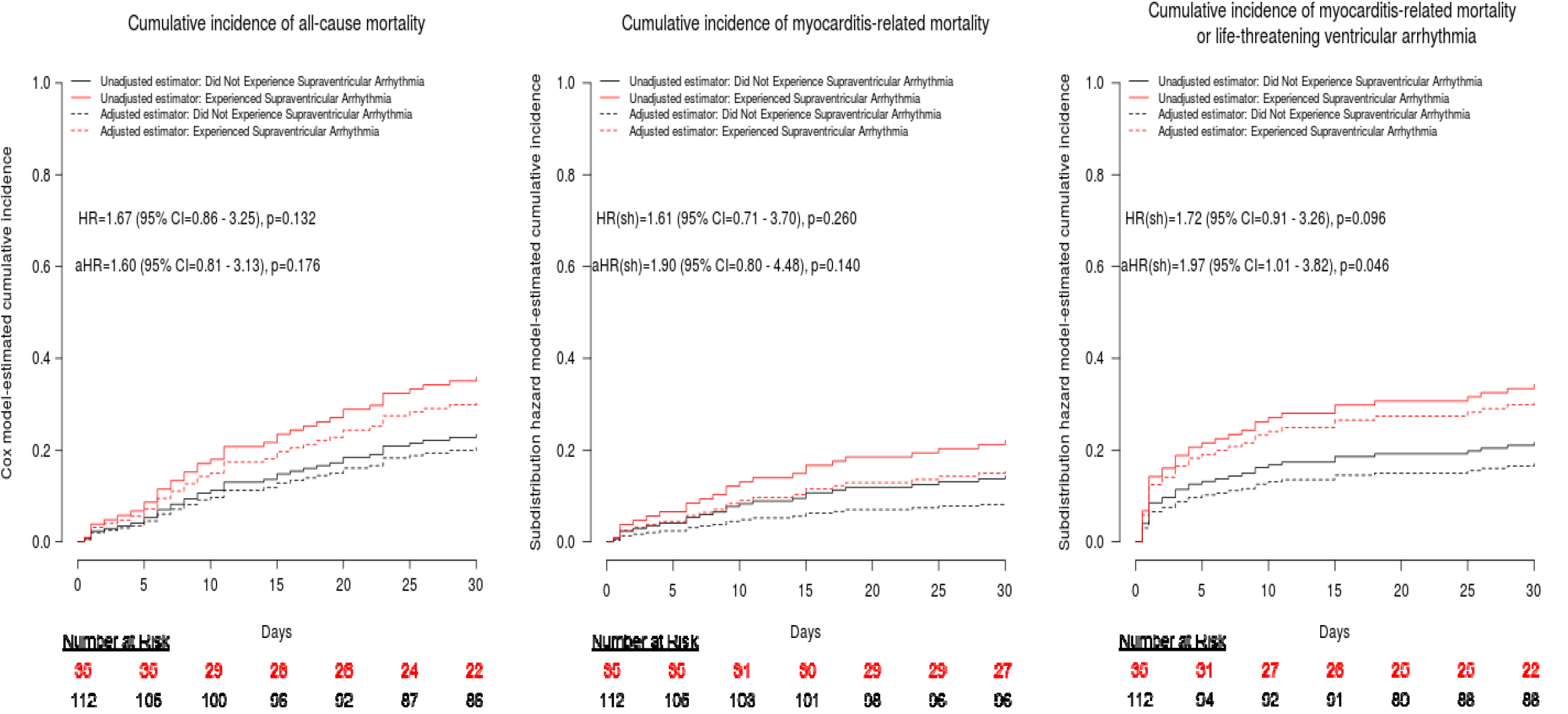
Outcomes by cumulative incidence of supraventricular arrhythmia.

**Supplemental Figure 3:**
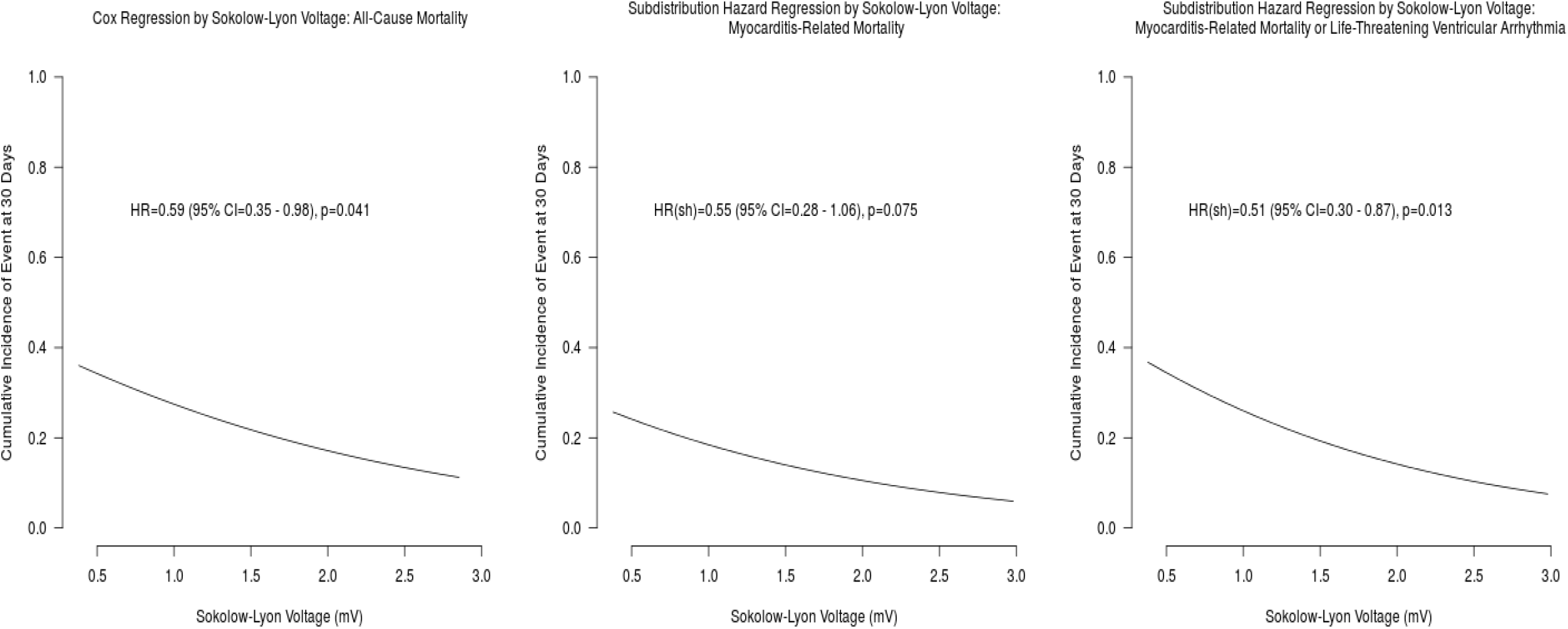
Model-estimated Cumulative Incidence of Event at 30-day by Sokolow-Lyon Index.

**Supplemental Figure 4:**
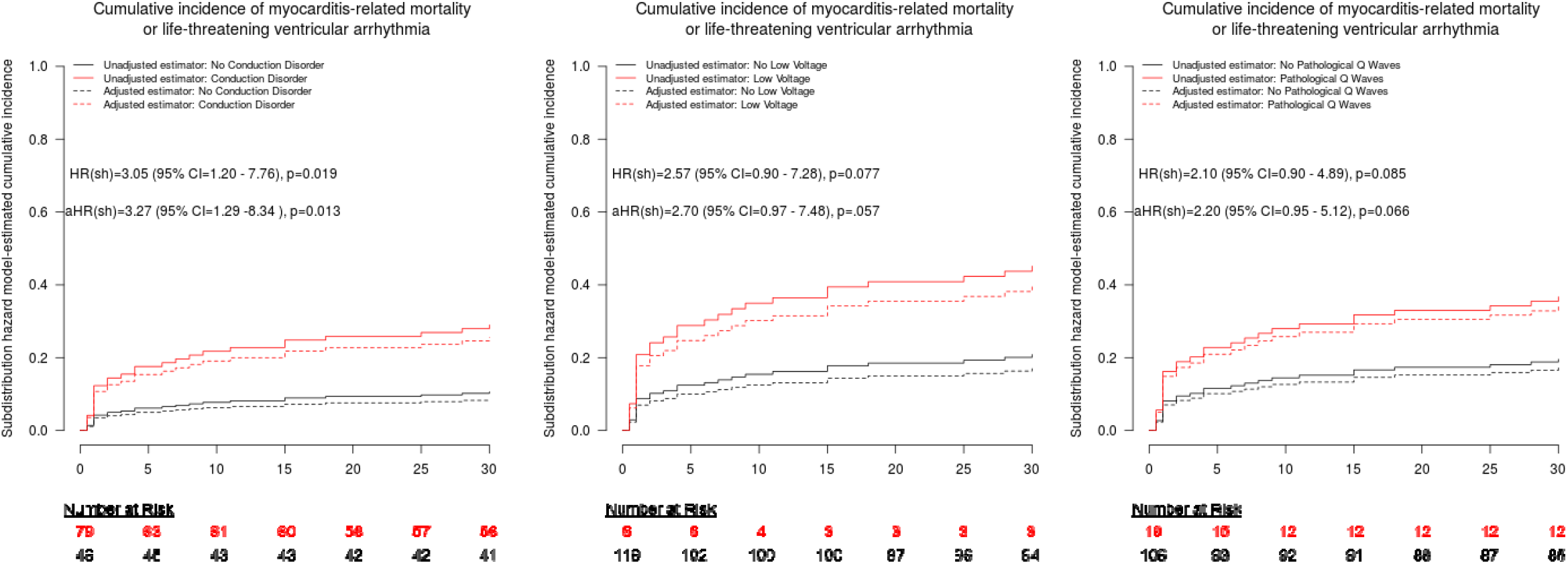
Cumulative incidence function by presenting ECG findings (composite outcome)

## References

1. Haslam A, Prasad V. Estimation of the Percentage of US Patients With Cancer Who Are Eligible for and Respond to Checkpoint Inhibitor Immunotherapy Drugs. JAMA network open. 2019;2(5):e192535–e192535. doi:10.1001/jamanetworkopen.2019.2535

2. Johnson DB, Balko JM, Compton ML, et al. Fulminant Myocarditis with Combination Immune Checkpoint Blockade. The New England journal of medicine. 2016;375(18):1749–1755. doi:10.1056/NEJMoa1609214

3. Champion SN, Stone JR. Immune checkpoint inhibitor associated myocarditis occurs in both high-grade and low-grade forms. Modern pathology□: an official journal of the United States and Canadian Academy of Pathology, Inc. 2020;33(1):99–108. doi:10.1038/s41379-019-0363-0

4. Salem J-E, Allenbach Y, Vozy A, et al. Abatacept for Severe Immune Checkpoint Inhibitor-Associated Myocarditis. The New England journal of medicine. 2019;380(24):2377–2379. doi:10.1056/NEJMc1901677

5. Salem J-E, Manouchehri A, Moey M, et al. Cardiovascular toxicities associated with immune checkpoint inhibitors: an observational, retrospective, pharmacovigilance study. The Lancet Oncology. 2018;19(12):1579–1589. doi:10.1016/S1470-2045(18)30608-9

6. Mahmood SS, Fradley MG, Cohen J V, et al. Myocarditis in Patients Treated With Immune Checkpoint Inhibitors. Journal of the American College of Cardiology. 2018;71(16):1755–1764. doi:10.1016/j.jacc.2018.02.037

7. Norwood TG, Westbrook BC, Johnson DB, et al. Smoldering myocarditis following immune checkpoint blockade. Journal for immunotherapy of cancer. 2017;5(1):91. doi:10.1186/s40425-017-0296-4

8. Zhang L, Awadalla M, Mahmood SS, et al. Cardiovascular magnetic resonance in immune checkpoint inhibitor-associated myocarditis. European heart journal. February 2020. doi:10.1093/eurheartj/ehaa051

9. Bonaca MP, Olenchock BA, Salem J-E, et al. Myocarditis in the Setting of Cancer Therapeutics: Proposed Case Definitions for Emerging Clinical Syndromes in Cardio-Oncology. Circulation. 2019;140(2):80–91. doi:10.1161/CIRCULATIONAHA.118.034497

10. Locke TJ, Karnik R, McGregor CG, Bexton RS. The value of the electrocardiogram in the diagnosis of acute rejection after orthotopic heart transplantation. Transplant international□: official journal of the European Society for Organ Transplantation. 1989;2(3):143–146. doi:10.1007/bf02414601

11. Kowalski O, Zakliczyński M, Lenarczyk R, et al. Electrophysiologic parameters suggesting significant acute cellular rejection of the transplanted heart. Annals of transplantation. 2006;11(1):35–39.

12. Geraud A, Gougis P, Vozy A, et al. Clinical Pharmacology and Interplay of Immune Checkpoint Agents: A Yin-Yang Balance. Annual review of pharmacology and toxicology. September 2020. doi:10.1146/annurev-pharmtox-022820-093805

13. Zhang L, Zlotoff DA, Awadalla M, et al. Major Adverse Cardiovascular Events and the Timing and Dose of Corticosteroids in Immune Checkpoint Inhibitor-Associated Myocarditis. Circulation. 2020;141(24):2031–2034. doi:10.1161/CIRCULATIONAHA.119.044703

14. Jain V, Mohebtash M, Rodrigo ME, Ruiz G, Atkins MB, Barac A. Autoimmune Myocarditis Caused by Immune Checkpoint Inhibitors Treated With Antithymocyte Globulin. Journal of immunotherapy (Hagerstown, Md□: 1997). 2018;41(7):332–335. doi:10.1097/CJI.0000000000000239

15. Tay RY, Blackley E, McLean C, et al. Successful use of equine anti-thymocyte globulin (ATGAM) for fulminant myocarditis secondary to nivolumab therapy. British journal of cancer. 2017;117(7):921–924. doi:10.1038/bjc.2017.253

16. Bonaros N, Dunkler D, Kocher A, et al. Ten-year follow-up of a prospective, randomized trial of BT563/bb10 versus anti-thymocyte globulin as induction therapy after heart transplantation. The Journal of heart and lung transplantation□: the official publication of the International Society for Heart Transplantation. 2006;25(9):1154–1163. doi:10.1016/j.healun.2006.03.024

17. Ruan V, Czer LSC, Awad M, et al. Use of Anti-Thymocyte Globulin for Induction Therapy in Cardiac Transplantation: A Review. Transplantation proceedings. 2017;49(2):253–259. doi:10.1016/j.transproceed.2016.11.034

18. Caforio ALP, Pankuweit S, Arbustini E, et al. Current state of knowledge on aetiology, diagnosis, management, and therapy of myocarditis: a position statement of the European Society of Cardiology Working Group on Myocardial and Pericardial Diseases. European heart journal. 2013;34(33):2636-2648, 2648a-2648d. doi:10.1093/eurheartj/eht210

19. Harris PA, Taylor R, Thielke R, Payne J, Gonzalez N, Conde JG. Research electronic data capture (REDCap)--a metadata-driven methodology and workflow process for providing translational research informatics support. Journal of biomedical informatics. 2009;42(2):377–381. doi:10.1016/j.jbi.2008.08.010

20. Harris PA, Taylor R, Minor BL, et al. The REDCap consortium: Building an international community of software platform partners. Journal of biomedical informatics. 2019;95:103208. doi:10.1016/j.jbi.2019.103208

21. Stewart S, Winters GL, Fishbein MC, et al. Revision of the 1990 working formulation for the standardization of nomenclature in the diagnosis of heart rejection. The Journal of heart and lung transplantation□: the official publication of the International Society for Heart Transplantation. 2005;24(11):1710–1720. doi:10.1016/j.healun.2005.03.019

22. Hickey KT, Sciacca RR, Chen B, et al. Electrocardiographic Correlates of Acute Allograft Rejection Among Heart Transplant Recipients. American journal of critical care□: an official publication, American Association of Critical-Care Nurses. 2018;27(2):145–150. doi:10.4037/ajcc2018862

23. Escudier M, Cautela J, Malissen N, et al. Clinical Features, Management, and Outcomes of Immune Checkpoint Inhibitor-Related Cardiotoxicity. Circulation. 2017;136(21):2085–2087. doi:10.1161/CIRCULATIONAHA.117.030571

24. Sandhu JS, Curtiss EI, Follansbee WP, Zerbe TR, Kormos RL. The scalar electrocardiogram of the orthotopic heart transplant recipient. American heart journal. 1990;119(4):917–923. doi:10.1016/s0002-8703(05)80332-1

25. Nakashima H, Katayama T, Ishizaki M, Takeno M, Honda Y, Yano K. Q wave and non-Q wave myocarditis with special reference to clinical significance. Japanese heart journal. 1998;39(6):763–774. doi:10.1536/ihj.39.763

26. Morgera T, Di Lenarda A, Dreas L, et al. Electrocardiography of myocarditis revisited: clinical and prognostic significance of electrocardiographic changes. American heart journal. 1992;124(2):455–467. doi:10.1016/0002-8703(92)90613-z

27. Keren A, Gillis AM, Freedman RA, et al. Heart transplant rejection monitored by signal-averaged electrocardiography in patients receiving cyclosporine. Circulation. 1984;70(3 Pt 2):I124–9.

28. Rassi AJ, Rassi A, Little WC, et al. Development and validation of a risk score for predicting death in Chagas’ heart disease. The New England journal of medicine. 2006;355(8):799–808. doi:10.1056/NEJMoa053241

29. Pereira Barretto AC, Mady C, Arteaga-Fernandez E, et al. Right ventricular endomyocardial biopsy in chronic Chagas’ disease. American heart journal. 1986;111(2):307–312. doi:10.1016/0002-8703(86)90144-4

30. Hulsmans M, Clauss S, Xiao L, et al. Macrophages Facilitate Electrical Conduction in the Heart. Cell. 2017;169(3):510-522.e20. doi:10.1016/j.cell.2017.03.050

31. Wei SC, Meijers WC, Axelrod ML, et al. A genetic mouse model recapitulates immune checkpoint inhibitor-associated myocarditis and supports a mechanism-based therapeutic intervention. Cancer discovery. November 2020. doi:10.1158/2159-8290.CD-20-0856

32. Natali LC, Maddukuri P, Lucariello R, et al. Significant arrhythmias associated with Interleukin-2 therapy. Journal of Clinical Oncology. 2005;23(16_suppl):2588. doi:10.1200/jco.2005.23.16_suppl.2588

33. Salem J-E, Ederhy S, Lebrun-Vignes B, Moslehi JJ. Cardiac Events Associated With Chimeric Antigen Receptor T-Cells (CAR-T): A VigiBase Perspective. Journal of the American College of Cardiology. 2020;75(19):2521–2523. doi:10.1016/j.jacc.2020.02.070

34. Lefebvre B, Kang Y, Smith AM, Frey N V, Carver JR, Scherrer-Crosbie M. Cardiovascular Effects of CAR T Cell Therapy: A Retrospective Study. JACC CardioOncology. 2020;2(2):193–203. doi:10.1016/j.jaccao.2020.04.012

35. Alvi RM, Frigault MJ, Fradley MG, et al. Cardiovascular Events Among Adults Treated With Chimeric Antigen Receptor T-Cells (CAR-T). Journal of the American College of Cardiology. 2019;74(25):3099–3108. doi:10.1016/j.jacc.2019.10.038

36. Surawicz B, Childers R, Deal BJ, et al. AHA/ACCF/HRS recommendations for the standardization and interpretation of the electrocardiogram: part III: intraventricular conduction disturbances: a scientific statement from the American Heart Association Electrocardiography and Arrhythmias Committee. Journal of the American College of Cardiology. 2009;53(11):976–981. doi:10.1016/j.jacc.2008.12.013

37. Thygesen K, Alpert JS, Jaffe AS, et al. Fourth Universal Definition of Myocardial Infarction (2018). Journal of the American College of Cardiology. 2018;72(18):2231–2264. doi:10.1016/j.jacc.2018.08.1038

38. Hancock EW, Deal BJ, Mirvis DM, et al. AHA/ACCF/HRS recommendations for the standardization and interpretation of the electrocardiogram: part V: electrocardiogram changes associated with cardiac chamber hypertrophy: a scientific statement from the American Heart Association Electrocardiograph. Journal of the American College of Cardiology. 2009;53(11):992–1002. doi:10.1016/j.jacc.2008.12.015

